# Elevated expression of *B4GALT6*, *GABRA1*, *GAD2*, *GLRA3*, *HTR2A*, *PCSK1*, and *SLC17A6* are postmortem markers for the ALS-Ox subtype

**DOI:** 10.1101/2024.03.21.24304538

**Authors:** Jarrett Eshima, Taylor R. Pennington, Raiyan Choudhury, Jordan M. Garcia, John Fricks, Barbara S. Smith

**Affiliations:** School of Biological and Health Systems Engineering, Arizona State University, Tempe, Arizona, 85287, USA; Trinity College of Arts & Sciences, Duke University, Durham, North Carolina, 27708, USA; School of Mathematical and Statistical Sciences, Arizona State University, Tempe, Arizona, 85287, USA

**Keywords:** Amyotrophic Lateral Sclerosis, patient stratification, subtypes, spinal cord, transcriptomics, concordance

## Abstract

In this work, we stratify 428 bulk RNA-seq spinal cord transcriptomes from 206 ALS patients to assess concordance with cortical phenotype. We find the postmortem spinal cord generally recaptures the molecular phenotypes observed in the postmortem cortex and observe weaker differences in patient survival after correction for repeat measures. We compare intra-patient subtype assigned in the cortex and spinal cord, finding modest agreement between the cortex and lumbar region specifically, ranging from 46.2 – 66.7%. We leverage differential expression analysis to identify seven marker genes for the oxidative stress (ALS-Ox) subtype that show consistent upregulation in both the cortex and spinal cord and utilize three to construct classifiers that achieve notable predictive power for the ALS-Ox subtype in three different holdout cohorts, with AUCs ranging from 0.81 – 0.89. Our study shows the ALS-Ox subtype is conserved in the spinal cord of the same patients, offering a postmortem foundation for clinical stratification.

## Introduction

Variability in the presentation and progression of Amyotrophic Lateral Sclerosis (ALS) has contributed to the lack of effective treatments, limited clinical trial success, and difficulty establishing reliable and neurodegenerative-specific biomarkers. Previous efforts to better understand patient heterogeneity utilize global gene expression profiles from the frontal and motor postmortem cortex and have demonstrated that distinct molecular states are present^1–3^. Using publicly available data^4^ (Gene Expression Omnibus Accession: GSE153960), our previous study identified three distinct molecular phenotypes in the ALS postmortem cortex and established an association between patient subtypes and variability in survival and age of onset^5^. We further clarify the presentation of subtypes in the ALS cohort by considering the existence of hybrid states, and show these phenotypes more closely reflect a continuous spectrum.

In this study, we expand our findings by considering the presentation and concordance of distinct molecular subtypes in the cervical, thoracic, and lumbar regions of the spinal cord from the same patients^4,5^. Using unsupervised clustering, we identify three distinct gene expression profiles and following enrichment, find these groups recapture the phenotypes presented in each cortical subtype. We then examine differences in patient clinical parameters, finding significant differences in survival by the majority agreement approach. However, after adjusting for repeat patient measures and other covariates using a Cox proportional hazard survival model, ALS subtype was not statistically significantly associated with survival. In fact, only two regions trended towards statistical significance (α < 0.1) using tissue-specific Cox models to ensure statistical independence. Considering agreement between the cortical and spinal subtypes within each patient, our results show intra-patient subtype agreement is modest yet statistically significant (34– 51%). We note that these results are partially affected by external covariates such as sequencing platform, site of collection, sex, and bulk tissue cell type composition^6^. The lumbar region shows the highest overall concordance with the frontal and motor cortex subtype in this cohort (43.4%; 161/371) and is least affected by location-dependent gene expression in the spinal cord. Both the cervical and thoracic regions appear inundated by cell type composition and other covariates, with >50% of all patient samples assigned the transcriptional dysregulation subtype (ALS-TD). Using differential expression, we find the ALS-Ox phenotype, defined by altered synaptic signaling and oxidative stress^1,5^, is highly conserved throughout the ALS central nervous system with region-independent elevated expression of transcripts *B4GALT6, GABRA1, GAD2, GLRA3, HTR2A, PCSK1,* and *SLC17A6* relative to other ALS subtypes and non-neurological controls. Lastly, we leverage these ALS-Ox marker genes to develop multiple classifiers that achieve promising stratification accuracies when applied to three holdout validation cohorts comprised of i) all cortex samples (*n*=585), ii) all spinal cord samples (*n*=519), and iii) all sample analyzed by HiSeq (*n*=415). Using RPKM-normalized expression of *B4GALT6*, *GLRA3*, *SLC17A6*, the average area under the ROC curve (AUC) was found to be ≥ 0.87 across all three validation cohorts, for multilayer perceptron and support vector machine classifiers. Comparable AUCs are observed when all seven marker genes were used to construct the classifiers, although small improvements are seen in the HiSeq validation cohort. Collectively our findings provide a roadmap to link cortical subtypes to the more accessible spinal cord, facilitating the implementation of patient subtyping to improve clinical trial design and therapeutic success.

## Results

### Clustering of spinal cord transcriptomes recaptures cortical phenotypes

To ascertain concordance between molecular subtypes presented in the ALS cortex with those presented in the spinal cord, we performed unsupervised clustering using 428 transcriptomes derived from cervical, thoracic and lumbar postmortem tissue, corresponding to 206 unique ALS patients (**Fig. S1; Table S1**). Transposable elements (TEs) were quantified for all patient samples using SQuIRE^7^, with transcript filtering criteria detailed previously^5^ (see Methods; **Supplemental Dataset 1**). Covariate-dependent gene expression was determined using DESeq2^8^ and all features differentially expressed due to (i) sex, (ii) site of collection, (iii) RIN, and (iv) tissue region were removed prior to clustering. Previous work shows differential gene expression in the ALS spinal cord is partially driven by cell type composition^6^. To help address bulk tissue effects during clustering, we further elected to remove oligodendrocyte, microglia, astrocyte, and endothelial cell marker genes (n = 1282), included in *Supplementary Table 3* from Humphrey *et al.*^6^. The majority of these features showed gene expression dependent on one of the four covariates previously addressed for both the NovaSeq (1061/1282) and HiSeq (855/1282) cohorts. We further address the sequencing platform covariate by splitting our cohort according to analytical platform (HiSeq 2500 and NovaSeq 6000, Illumina, San Diego, CA) and running the stratification analyses separately^5^. Factorization rank was estimated from clustering metrics (**Fig. S2**), and a rank of three was used in both cohorts.

After applying a variance stabilizing transformation (VST)^8^, the top 5000 most variably expressed transcripts were selected for non-smooth non-negative matrix factorization (nsNMF)^9^ using SAKE^10^. Our results capture three distinct groups in both the NovaSeq and HiSeq cohorts. These groups show distinct expression profiles (**Fig. 1A, 1B**) and yield three clusters following application of principal component analysis (**Fig. 1C, 1D**). To better understand biological context of these stratified groups, we performed enrichment using all 5000 features from the NovaSeq and HiSeq cohorts, and combined them to yield 8163 non-duplicate transcripts (corresponding to 5438 gene symbols). The resulting feature set was used to perform hypergeometric enrichment analysis with Enrichr^11^ and the Reactome^12^ pathway database, according to our previously described approach^5^. We further leveraged GSEA^13^ to enrich each stratified group against a non-neurological control cohort comprised of 91 donor samples from the cervical, thoracic, and lumbar regions of the spinal cord (**Fig. 1E, 1F; Table S1**). In agreement with the phenotypes identified in the ALS postmortem cortex^5^, we observe significant enrichment for neuroinflammatory signatures in ALS-Glia patients when compared to the other two subtypes. Interestingly, normalized enrichment scores indicate negative enrichment for neuroinflammatory pathways in the ALS spinal cord, relative to controls, whereas positive enrichment is observed in the cortex^5^ – findings which may be linked to cell type composition^6^ and our stringent filtering of glial marker genes. In ALS-Ox patients, we observe statistically significant positive enrichment for genes associated with synaptic signaling, mirroring the phenotype observed in the postmortem cortex. Pathways associated with RNA metabolism and processing were the most strongly enriched in the ALS-TD subtype when compared to controls using GSEA, although significant associations were not observed at an adjusted *p*-value < 0.05 by either enrichment approach. Collectively, our findings demonstrate that unsupervised clustering of the ALS spinal cord recapitulates many of the same subtype signatures observed in the postmortem cortex. Conversely, phenotypic differences between the groups are less pronounced than in the cortex, evident in the magnitude of GSEA normalized enrichment scores. These findings likely reflect differences in cell type composition between the two regions of the central nervous system and the fact that most glial marker genes demonstrated covariate-dependent expression and were subsequently filtered.

**Fig. 1.**
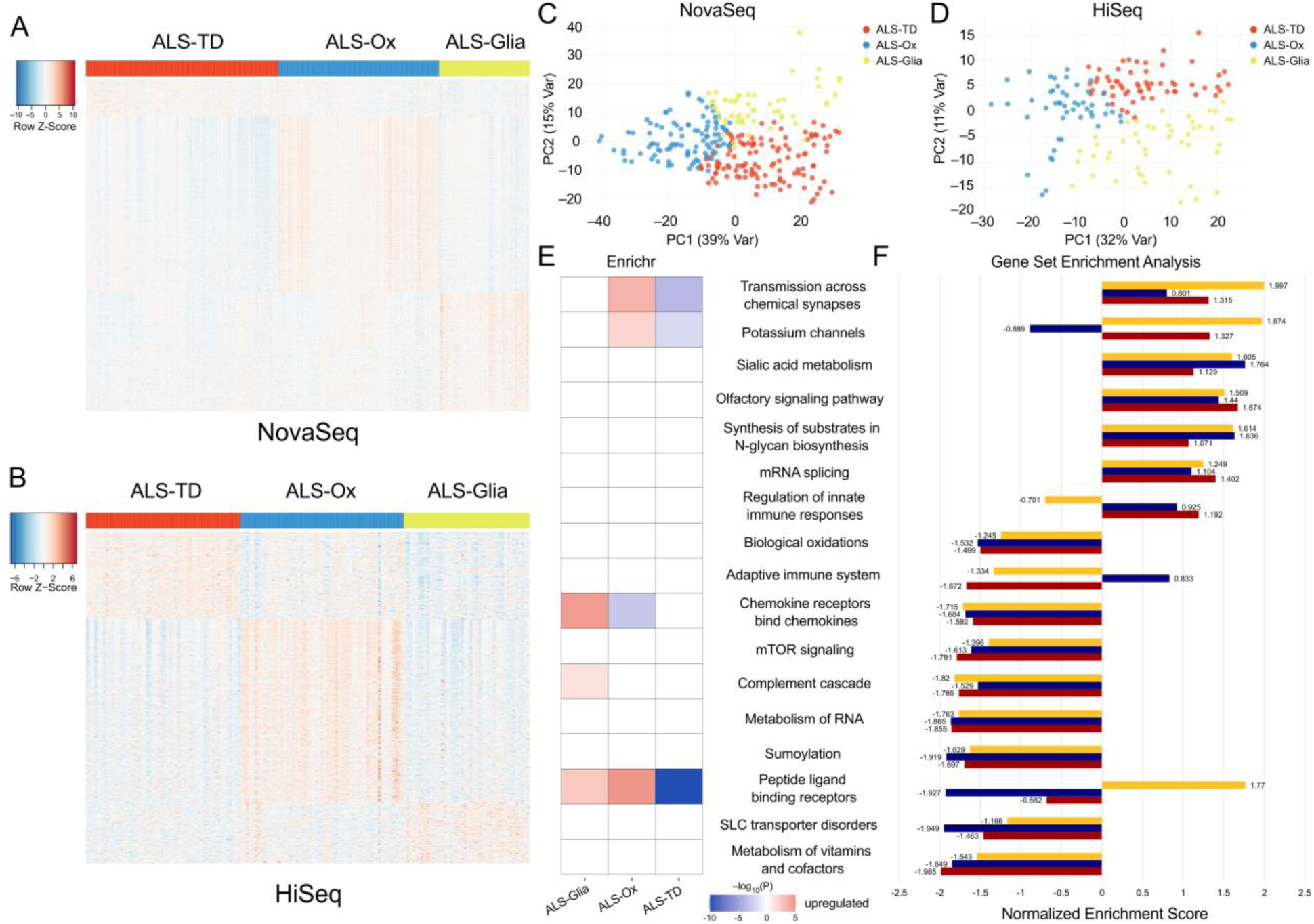
Clustering and enrichment of ALS spinal cord transcriptomes. (A) Heatmap showing subtype specific gene expression in the NovaSeq cohort, comprised of 273 biologically independent tissue samples and 763 transcripts. (B) Gene expression in the HiSeq cohort, with 155 biologically independent tissue samples and 567 transcripts. Both heatmaps are presented after z-score normalization with features selected by SAKE^10^. Principal component analysis shows three distinct clusters when considering the first two principal components, in both the (C) NovaSeq and (D) HiSeq cohorts. Enrichment analysis using (E) Enrichr Fisher exact tests with FDR^15^ adjusted *p*-values presented on the –log_10_ scale. Genes were assigned to patient clusters depending on median expression and downregulated pathways are indicated with the negative magnitude of the –log_10_ transformed *p*-value. (F) Gene set enrichment analysis (GSEA) with non-neurological controls specified as the reference level for calculation of all normalized enrichment scores.

Allocation to each of the three subtypes in the NovaSeq cohort more closely agrees with findings from the frontal and motor cortex^5^, with ALS-Glia representing the rarest subtype (21.9% of spinal transcriptomes compared to 19.2% in the cortex^5^) corresponding to a Glia:Ox:TD ratio of 1:1.5:2 (**Fig. S3**). In the HiSeq cohort, a Glia:Ox:TD subtype ratio of 1:1:1.4 was observed, corresponding to ∼30% of patients classified as ALS-Glia, indicating the selected sequencing platform influences the detectable subtype expression signature (**Fig. S3**). In both cases, the transcriptional dysregulation (TD) subtype was the most commonly assigned – as compared to the oxidative stress subtype in the postmortem cortex^5^ – potentially a consequence of weak dependency on the removed covariates (**Fig. S4A-D**), weak neuronal expression associated with cell type composition in the spinal cord (**Fig. S4E**), and stringent filtering of covariate-dependent genes. Patient- and sample-level phenotype information, including subtype is provided in **Supplemental Dataset 2**.

### Differences in subtype survival are primarily driven by repeat patient measures

After stratification of the spinal cord cohort, we examined patient clinical parameters to determine if subtype level differences in survival are maintained. A Kaplan-Meier survival analysis^14^ was performed after assigning patient-level subtype using majority agreement between all available regions of the spinal cord or if a single tissue sample was characterized for a given patient (26/206; 12.6%). Similar to the postmortem cortex^5^, we observe a significantly shorter survival duration in the ALS-Glia subtype when compared to ALS-Ox (*p* = 0.032) and Discordant (*p* = 0.023) groups but not the ALS-TD subtype (*p* = 0.27) (**Fig. S5A**). The higher proportion of glial cells comprising spinal cord tissue (**Fig. S4E**) likely drives weaker phenotypic differences between ALS-Glia and ALS-TD subtypes, relative to the cortex^5^, which may partially explain the similar survival curves observed. The mean survival duration for ALS-Glia patients was found to be 31.3 ± 3.97 months (mean ± SE), while ALS-Ox patients were found to have the longest mean survival duration at 45.6 ± 4.87 months (**Table S1**). Interestingly, we observe a significant difference in the age of disease onset between ALS-Glia and discordant patients after a false discovery rate^15^ (FDR) correction (FDR *p* = 0.018), strengthening the trend seen in the postmortem cortex^5^ (**Fig. S5B**). Differences in age at death were not significant after FDR correction (**Fig. S5C**). No significant relationships were found between disease comorbidity and ALS subtype in the spinal cord using Chi-squared tests of independence (**Fig. S5D, S5E**). Collectively, findings generally agree with results from the postmortem cortex^5^, but also reflect some of the challenges associated with stratifying patients using spinal cord gene expression.

To better understand how repeat patient measures and the majority agreement approach^1,5^ influences previously observed survival differences in the cortex^5^, we performed multivariate survival analyses independently in each tissue region using Cox proportional hazard regression^16–20^. Survival analysis in each region of the CNS shows weaker differences in disease duration dependent on subtype, revealing that sex and disease group (ALS-FTLD, ALS/Alzheimer’s, ALS-SOD1, and ALS-TDP) contribute more to differences in disease duration (**Fig. 2**). Model diagnostics show the proportional hazard assumption is met for all covariates (**Fig. S6**). Reference levels are set as males, ALS-TDP disease group, and the ALS-TD subtype. Subtype effect on survival was not statistically significant in the six regions presented, although the ALS-Ox subtype trended towards significance in the cervical (*p* = 0.095) and thoracic (*p* = 0.064) tissues with hazard ratios between 0.5 and 0.9. The ALS-Glia covariate is less consistent, with hazards ranging from 0.77 to 1.4, but were more generally equivalent or slightly higher than the ALS-TD reference level. Large confidence intervals seen in the disease group covariate likely reflect low sample number and/or naturally higher variability, although it is notable to see the greatest difference in hazard ratio in this covariate and shows disease comorbidity negatively affects risk of death. Interestingly, sex-dependent differences in hazard ratio are seen, with females showing an increased risk for death in the range of 14–80% for the six regions presented. We further consider the same model, excluding the disease group covariate to account for possible collinearity with subtype. We find similar results with the ALS-Ox hazard ratio between 0.58 and 0.93, ALS-Glia hazard ratio between 0.82 and 1.57, and elevated hazard in females (1.10 – 1.81). Taken together, the ALS-Ox subtype may be weakly associated with a better patient prognosis, however the inclusion sex, age, and disease group covariates while maintaining observational independence demonstrates limitations with the majority agreement approach and that the effects due to these factors contributes more to survival differences than subtype. The phenotype information used to construct the Cox proportional hazard model is provided in **Supplemental Dataset 3.**

**Fig. 2.**
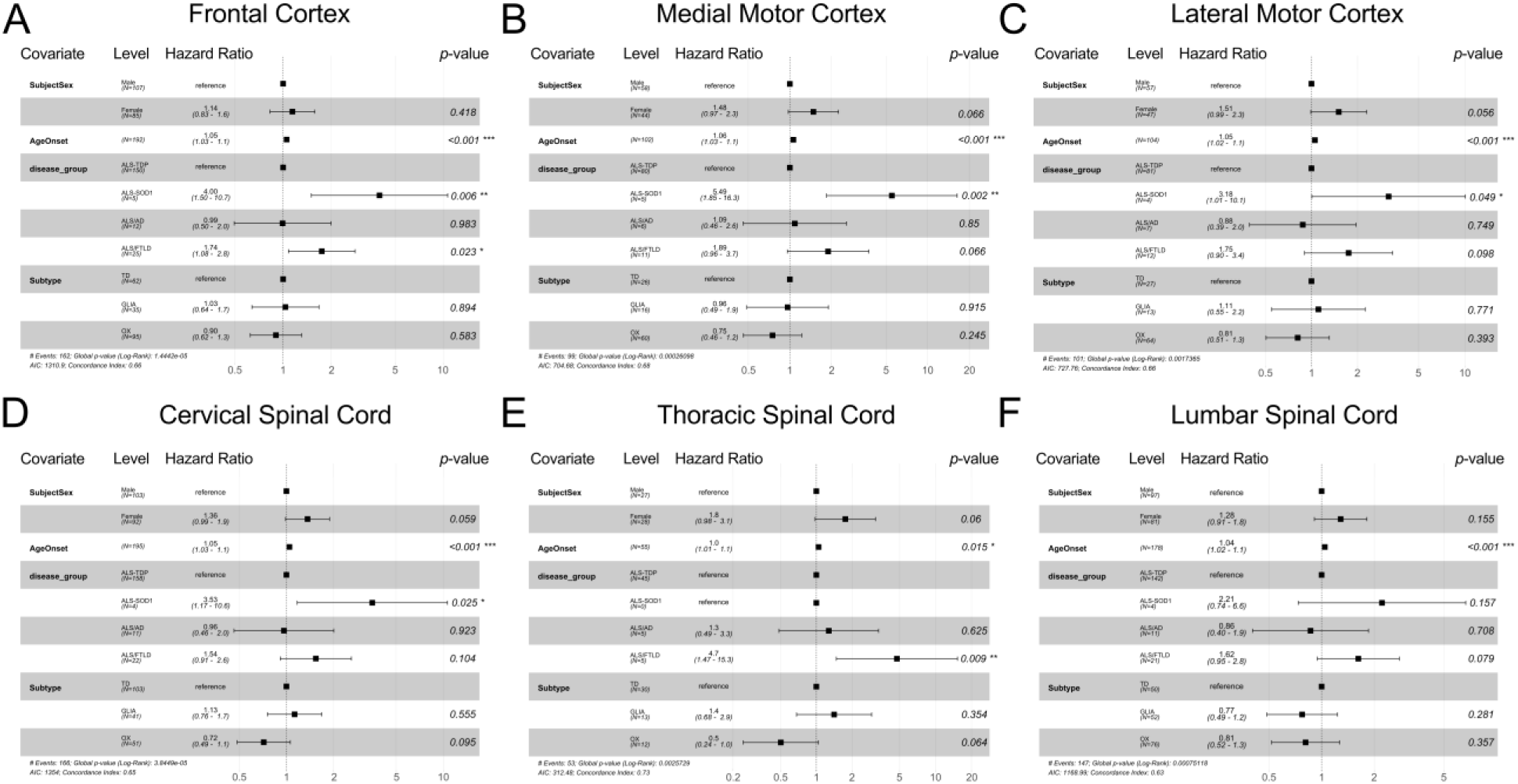
Independent and Identically Distributed (IID) Survival Analysis. Tissue region specific survival analyses for the ALS (A) frontal cortex (n = 193), (B) medial motor cortex (n = 102), (C) lateral motor cortex (n = 104), (D) cervical spinal cord (n = 195), (E) thoracic spinal cord (n = 55), and (F) lumbar spinal cord (n = 178). The “unspecified motor cortex” (n = 52) was not considered. The effects due to sex and disease group can be seen to contribute more to survival differences among patients however, across most tissue regions, there is a nonsignificant but consistent trend toward a lower hazard associated with the ALS-Ox subtype. Model terms are presented as hazard ratios with the 95% confidence interval shown. Terms are separated by covariate and subgroup, with reference levels indicated.

### Concordance between cortical and spinal subtypes is dependent on tissue and sequencing platform

Given the high degree of overlap between the patients considered in our previous study^5^ and those included in this analysis (**Fig. S1B**), we worked to better understand the agreement between the postmortem cortex (frontal, lateral motor, medial motor, and ‘unspecified’ motor regions) and spinal cord in presentation of ALS subtype. Concordance was considered at the sample level, to ensure independence, and presented as a matrix of pie charts, with spinal cord region along the rows and cortical region along the columns (**Fig. 3**). Excluding the unspecified motor cortex samples, we find the highest concordance between the frontal cortex and lumbar region of the spinal cord for the ALS-Glia subtype (46.2%), the medial motor cortex and lumbar region for the ALS-Ox subtype (46.4%), and the medial motor cortex and cervical spinal cord for the ALS-TD subtype (69.6%). The higher concordance observed in the ALS-TD subtype, and generally lower overall agreement, likely stems from bias towards the ALS-TD subtype during unsupervised clustering, despite removal of covariate dependent genes (**Fig. S4A-D**), and differences in cell type composition as compared to the cortex^5^ (**Fig. S4E**). Similarly, the lumbar region is seen to more closely reflect the proportion of subtypes observed in the cortex^5^ (**Fig. S4C**), generally corresponding to a noticeable improvement in concordance between the cortex and lumbar spinal cord, relative to other spinal cord regions, for ALS-Ox and ALS-Glia patients (**Fig. 3**). More generally, concordance between the lumbar spinal cord and each region of the cortex, excluding the lateral motor, is statistically significant with agreement being higher than would be expected by random chance. Using bootstrapping, distributions for the expected number of concordant patient samples were generated from 10,000 iterations, after adjusting sampling probabilities to reflect subtype proportions observed in the cortex and spinal cord. True concordant values were compared against the derived distribution for estimation of *p*-values, assuming a one-tailed binomial distribution, which were found to be 0.012 in the frontal cortex, 0.031 in the medial motor cortex, 0.160 in the lateral motor cortex, and 0.021 in the ‘unspecified’ motor cortex. Statistically significant concordance was also observed between the cervical region of the spinal cord and the frontal cortex (*p* = 0.026).

**Fig. 3.**
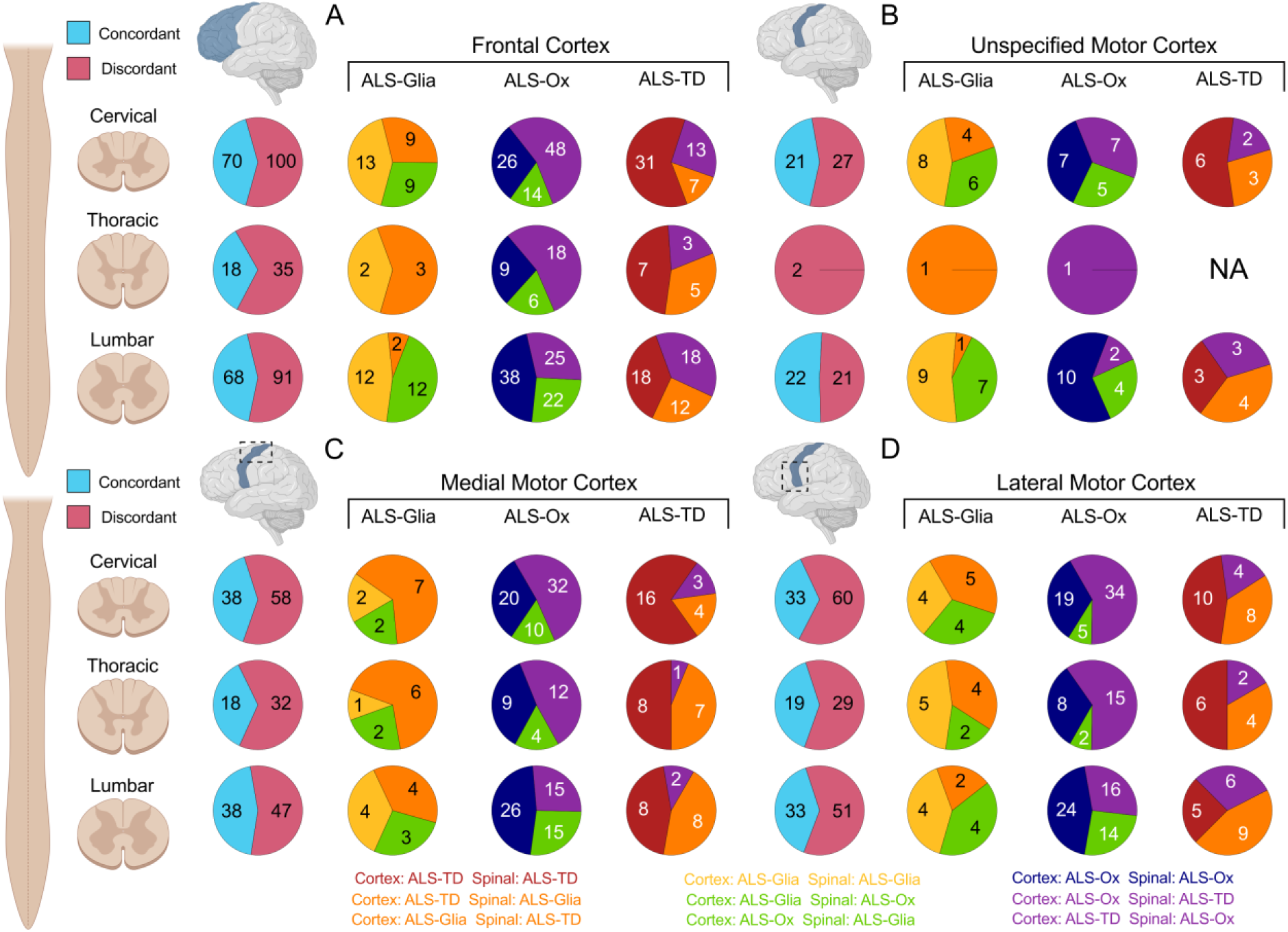
Subtype concordance between the ALS cortex and spinal cord. Agreement between the subtype assigned to the cervical, thoracic, and lumbar regions (*rows*) and (A) the frontal cortex, (B) the unspecified motor cortex, (C) the medial motor cortex, and (D) the lateral motor cortex – for all available samples. Pie charts are first presented as an aggregate of all paired tissue samples (light blue and pink) and in a subtype-specific manner. Concordance at the subtype level (*columns*) has been color coded to indicate agreement (gold, navy, and maroon) or disagreement (orange, green, and purple) between the two tissue regions compared. No patients assigned ALS-TD in the unspecified motor cortex had a corresponding thoracic spinal cord sample in this cohort. Created with BioRender.com.

We extended our concordance analysis by further separating patients by sequencing platform to assess dependence on instrumentation (**Fig. S3, S7, S8, S9**). We find the NovaSeq 6000 sequencing platform outperforms the HiSeq 2500 sequencing platform in the cervical (42.3% vs 35.4%) and lumbar spinal cord (47.7% vs 35.4%) but not the thoracic spinal cord (32.0% vs 37.9%). In the NovaSeq cohort, the highest concordance for the ALS-Glia subtype remains the same tissue pairing at 48.0%, and for ALS-Ox the highest agreement was seen between the lateral motor cortex and lumbar spinal cord at 48.4% (excluding the unspecified motor cortex and pairings with a single observation) – although lower sample numbers may partially explain these differences. In the NovaSeq subset, intra-patient concordance between the postmortem cortex and lumbar region of the spinal cord remains statistically significant in the frontal cortex (*p* = 0.024), medial motor cortex (*p* = 0.041), lateral motor cortex (*p* = 0.037), and ‘unspecified’ motor cortex (*p* = 0.010). Additionally, concordance was significant between the cervical region of the spinal cord and the frontal cortex (*p* = 0.024) and ‘unspecified’ motor cortex (*p* = 0.050). No tissue pairings were found to be statistically significant in the HiSeq subset. Collectively our analysis shows weak to moderate agreement in subtype presented throughout the ALS postmortem cortex and spinal cord, despite differences in cell type composition, after removal of covariate-dependent genes. We further demonstrate concordance between the cortex and spinal cord phenotype is dependent on tissue and sequencing platform, and find the lumbar region of the spinal cord shows the highest overall concordance with the cortical phenotype in this cohort. Importantly, bootstrapping shows that the proportion of concordant patients in the NovaSeq cohort consistently exceeds the number expected by random chance when comparing any region of the cortex with the lumbar region of the spinal cord, improving the likelihood that patient subtypes are expressed throughout the individual’s central nervous system and influence the disease course.

We further considered patient concordance by screening for patients assigned the same subtype in every sample considered in this study and the previous^5^. We find a total of 45 patients pass this criterion (45/222; 20.3%) (**Supplemental Dataset 4**) and filter these patients further to find 5 ALS-Glia patients, 12 ALS-Ox patients, and 19 ALS-TD patients coherently assigned a single subtype in *both* the cortex and spinal cord (**Fig. S10**). While the lower patient number limits the extrapolation of these findings, we observe a surprising association with sex and a stark difference in disease duration in this concordant patient subset (**Fig. S10**). Despite the statistical limitations of assigning patient subtype using the majority agreement approach, concordance between the majority postmortem cortex subtype^5^ and majority spinal cord subtype are considered using this method. We find that more than half of patients are discordant (68.2%), which likely reflects differences in cell type composition between the cortex and spinal cord and highlights the challenges of linking patient phenotype between these two regions (**Fig. S11**).

### Differential expression reveals transcripts *B4GALT6*, *GABRA1, GAD2, GLRA3, HTR2A, PCSK1*, and *SLC17A6* are elevated in the ALS-Ox cortex and spinal cord

We then applied differential expression^8^ to identify subtype-specific transcript expression in the full spinal cord cohort. After adjusting for sex, site of collection, RIN, tissue, and sequencing platform covariates, we find transcript expression that uniquely defines each subtype regardless of analytical platform (**Fig. 4A**). Differential expression *p*-values, after FDR adjustment and –log10 transformation, are presented as heatmaps using pairwise comparisons for all group combinations (**Fig. 4B**). All differential expression results are provided in **Supplemental Dataset 5**. Expression of transcripts stratifying ALS-Glia and ALS-TD subtypes in the spinal cord is weaker, evident in the heatmap and the differential expression *p*-values relative to the cortex^5^, and likely reflects differences in cell type composition in the spinal cord relative to the cortex^5^ (**Fig. S4E**). Most notably, we identify a total of ten transcripts with consistently elevated expression, irrespective of tissue region in the postmortem central nervous system, relative to the other subtypes and non-neurological controls in this cohort (**Fig. 4C, S12**). Seven of these transcripts were specific for the ALS-Ox subtype, while the remaining three were specific for ALS-Glia. A total of 1,104 unique transcriptomes were considered, from 5 distinct regions of the central nervous system, corresponding to 222 ALS patients, 88 non-neurological controls, and 42 frontotemporal dementia (FTLD) patients. ALS-Ox marker genes *GABRA1, GAD2, GLRA3, HTR2A, PCSK1,* and *SLC17A6* collectively implicate changes to synaptic signaling, with elevation of inhibitory receptors and enzymes involved in the biosynthesis of inhibitory neurotransmitters. Upregulation of *ST6GALNAC2* in ALS-Glia samples and *B4GALT6* in ALS-Ox suggests protein glycosylation may play a surprising role in the progression of ALS phenotype. As may be expected, expression of the subtype marker genes was generally different in the cortex and spinal cord regions. Notably, we find these genes better stratify this patient cohort when considering spinal cord expression, evident in the FDR-adjusted *p*-values (**Fig. 4C, S12**), which offers promise for clinical translation. Yet, difficulty stratifying ALS-Glia and ALS-TD patients in the spinal cord may limit the practicality of patient subtyping using Glia marker transcripts.

**Fig. 4.**
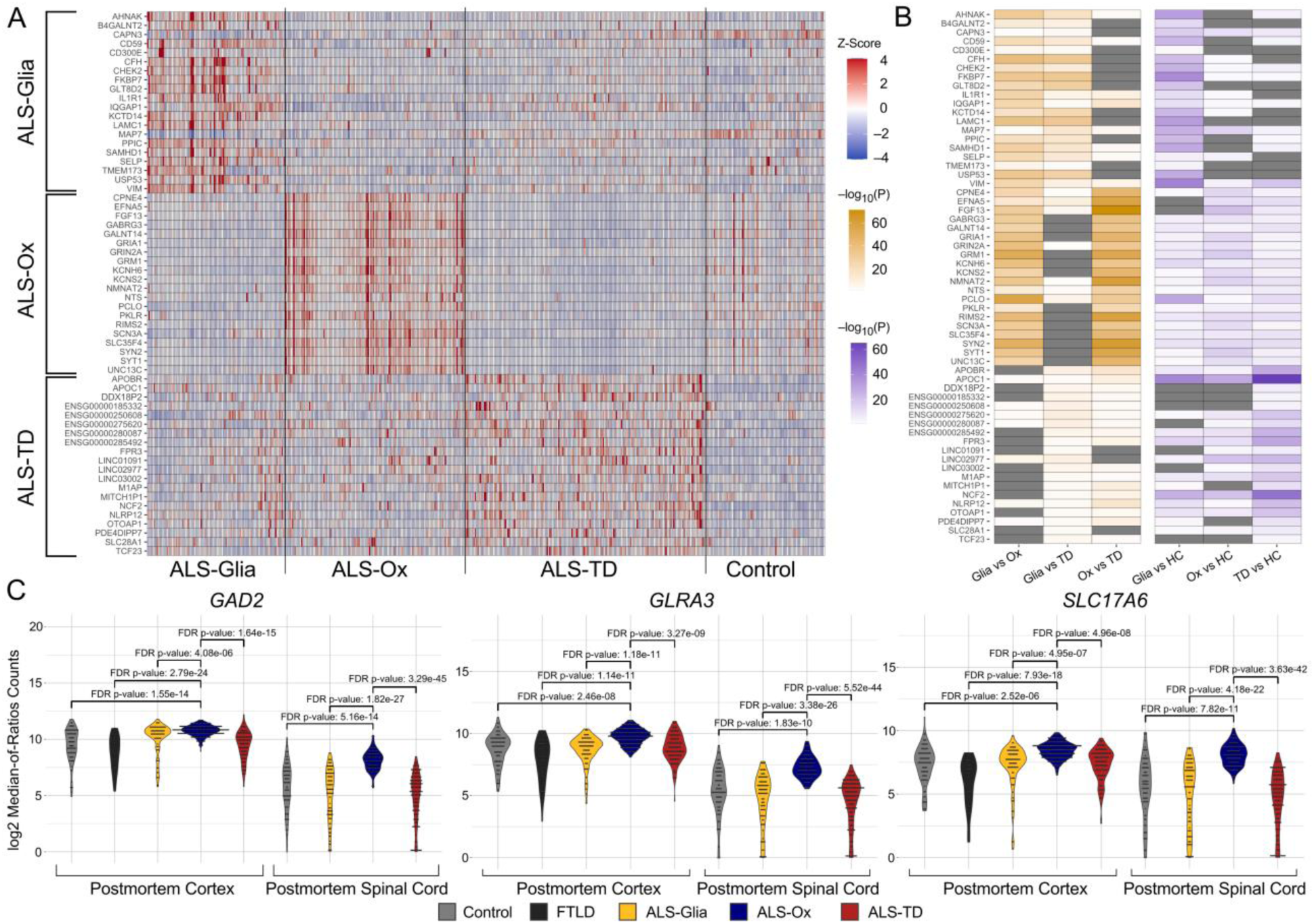
Differential expression identifies transcripts in the spinal cord that stratify subtypes and ALS-Ox markers shared between the cortex and spinal cord. (A) Heatmap showing z-score normalized expression following transformation to the median-of-ratios scale for each subtype. For plotting, z-scores < –4 or > 4 are adjusted to –4 and 4, respectively. All presented genes have mean raw counts > 10 and are expressed uniquely in a single ALS subtype. A total of 519 spinal cord samples are shown along the columns, grouped by subtype. (B) Heatmaps showing –log_10_ transformed differential expression FDR-adjusted *p*-values using pairwise comparisons. Gray cells indicated an adjusted *p*-value > 0.05. (C) Representative ALS-Ox marker transcripts, with expression shown throughout the postmortem cortex (frontal and motor) and spinal cord (cervical, thoracic, lumbar). Expression of *GABRA1*, *GAD2*, and *SLC17A6* are consistently and significantly elevated in ALS-Ox patients as compared to other subtypes and controls from comparable tissue regions. A combined total of 1,104 ALS and control samples are considered across all tissue regions presented. Using DESeq2^8^, expression normalization was performed independently for the cortex and spinal cord. In both cases, effects due to sex, sequencing platform, RIN, and site of collection covariates are captured in the design equation – while the tissue source covariate was uniquely included during normalization in the spinal cord cohort.

To demonstrate differential gene expression is not dependent on median-of-ratios transcript normalization, we further consider marker gene expression using RPKM normalization in a refined set of ALS patients with observations available from both the postmortem cortex and spinal cord (*n* = 192 ALS patients, 88 non-neurological controls) (**Fig. S13; Supplemental Datasets 2 and 6**). ALS patient samples were binned into one of three categories in an effort to capture the spectrum of phenotypes typically observed in most patients, which include: “Concordant ALS-Ox” (100% of tissue samples are ALS-Ox), “At least 50% ALS-Ox”, and “Generally not ALS-Ox” (<50% of tissue samples are ALS-Ox). All ALS-Ox marker genes show a decreasing trend in the median expression as intra-patient concordance for the ALS-Ox subtype decreases. Further, patients that generally don’t present as ALS-Ox maintain marker gene expression at a level similar to non-neurological controls. Collectively, these findings show that ALS-Ox marker genes established in this work provide a foundation to stratify ALS patients and account for the moderate intra-patient concordance observed between the cortex and spinal cord.

We extend our differential expression analysis by considering other relevant transcripts, including those found to stratify this cohort using postmortem cortex transcriptomes^5^. In the ALS-Ox spinal cord, we observe statistically significant upregulation of the *STMN2* transcript and the truncated pathological form associated with TDP-43 cryptic exon splicing when compared to the ALS-TD subtype, previously determined by Prudencio *et al.*^4^ (**Fig. S14A-B, Supplemental Dataset 2**). No significant differences in *TARDBP*, encoding TDP-43, were observed between the ALS subtypes in either the postmortem cortex or the spinal cord (**Fig. S14C**). Neuroinflammatory genes *AIF1*, *CD68*, *HLA-DRA*, *TREM2*, and *TYROBP* were among the most elevated transcripts in the cortex of ALS-Glia patients^5^ but not the spinal cord, likely reflecting regional differences in cell type populations (**Fig. S4E, S15**). Further these transcripts were included in the 1282 glial marker genes removed prior to clustering – which may partially explain the similar expression of these transcripts in ALS-Glia and ALS-TD subtypes. Oxidative and proteotoxic stress genes *BECN1*, *OXR1*, *SERPINI1*, *SOD1*, and *UBQLN2* generally show weaker differences in spinal cord expression when compared to the other two subtypes (**Fig. S15**). Similarly, transcriptional regulators miR24-2 and *NKX6-2* show specificity for the postmortem cortex, although *NKX6-2* expression is most elevated in the spinal cord of ALS-TD patients, relative to the other two subtypes (**Fig. S15**).

### Translation of ALS-Ox marker genes

Building off results from differential expression, we then utilize ALS-Ox marker genes *B4GALT6, GABRA1, GAD2, GLRA3, HTR2A, PCSK1,* and *SLC17A6* (**Fig. 4C, Fig. S12**) to develop multiple classifiers of varying complexity (**Fig. 5; Fig. S16, S17**). In each case, we assessed classifier performance using RPKM normalized expression, an 80/20 train-test split, 100-fold cross validation, two classes (“Ox” and “NotOx”), and three different holdout (validation) cohorts comprised of all (i) postmortem spinal cord samples (ii) postmortem cortex samples and (iii) HiSeq samples. The first holdout cohort estimates predictive accuracy when assigning spinal cord subtype using cortex expression, while the opposite is true in the second holdout cohort. The final holdout cohort is designed to better estimate predictive accuracy when applied to new patient cohorts accounting for instrument-dependent expression. While it may be reasonable to assume that predicting the cortex phenotype using spinal cord expression is more clinically useful as it limits diagnostic invasiveness, we construct models for both cases to demonstrate the capability of our subtype-specific transcripts to stratify the cohort regardless of region-dependent expression differences. All expression matrices and sample labels used to construct ALS-Ox classifiers are provided in **Supplemental Dataset 6**.

**Figure 5.**
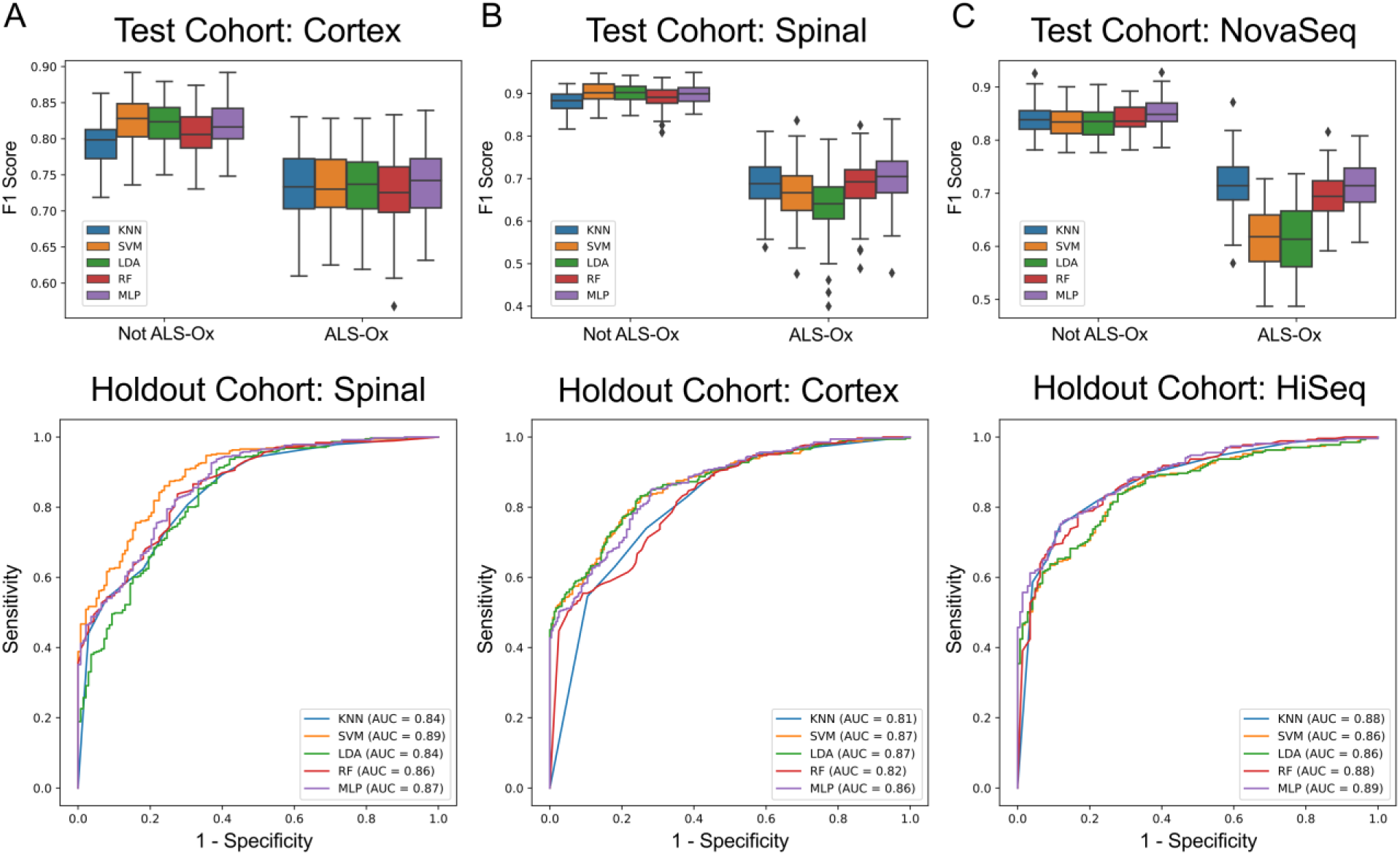
Supervised classification of ALS-Ox samples using expression of *B4GALT6*, *GLRA3*, and *SLC17A6* marker genes. Five different classification algorithms were considered. F1 scores obtained from 100-fold cross validation in the test cohort are presented first and separated by classification level (‘Ox’ vs ‘NotOx’). A combined total of 1,104 ALS and control samples are considered, with *n*=377 (∼34%) assigned the ALS-Ox label. The five classifiers were constructed and applied to three different holdout/validation cohorts comprised of (A) all postmortem spinal cord samples (*n*=519), (B) all postmortem cortex samples (*n*=585), and (C) all samples analyzed by HiSeq (*n*=415). ROC plots are presented second, for each classifier, and show sensitivity and 1-specificity metrics when applied to the specified holdout cohort.

With the aim of reducing clinical diagnostic burden, we constructed classifiers from all three-gene combinations of ALS-Ox marker genes and screened for predictive power using partial least squares discriminate analysis^21^ (PLS-DA). After training and testing our classifiers, we find the three-gene combination of *GAD2, GLRA3,* and *SLC17A6* slightly outperforms other gene combinations when predicting subtype in the spinal cord validation cohort (AUC = 0.927) (**Fig. S16A**). Conversely, when training on the spinal cord cohort, *HTR2A, SLC17A6,* and *B4GALT6* gene set showed the highest predictive accuracy after application to the cortex validation cohort (AUC = 0.881) (**Fig. S16B**). In the HiSeq validation cohort, *B4GALT6, GLRA3,* and *SLC17A6* demonstrated the highest predictive accuracy (AUC = 0.831), suggesting these transcripts may be more invariant to differences in sample preparation and instrumentation (**Fig. S16C**). Moreover, the same gene combination of *B4GALT6, GLRA3,* and *SLC17A6* demonstrated the highest average AUC across all three validation cohorts (AUC = 0.873), indicating these genes may be most robust for assigning ALS-Ox patient subtype. When compared to the PLS-DA classifier using all seven ALS-Ox marker genes, a decrease in predictive power is observed in the spinal cord holdout (AUC = 0.922), the cortex holdout (AUC = 0.861), and the HiSeq holdout (AUC = 0.809).

Building on promising results from PLS-DA, we extend our analysis by performing supervised machine learning using k-nearest neighbor (KNN), linear discriminant analysis (LDA), random forest (RF), support vector machine classifier (SVM), and multilayer perceptron (MLP) classification frameworks^22^. Classifiers were constructed using RPKM normalized expression of (i) the best three-gene combination from PLS-DA (*B4GALT6*, *GLRA3*, *SLC17A6*) (**Fig. 5**) and (ii) all seven ALS-Ox marker genes (**Fig. S17**). Using the top three discriminatory genes, the SVM classifier demonstrates the highest overall predictive accuracy when stratifying ALS-Ox and “not ALS-Ox” with median F1 scores from the test cohort ranging between 0.62–0.73 for ALS-Ox and 0.83–0.90 for ‘not ALS-Ox’, and holdout cohort AUCs ranging from 0.86–0.89 (**Fig. 5**). Similar performance is observed in the MLP classifier. In agreement with results observed during PLS-DA, the seven-gene classifier generally demonstrated worse predictive accuracy in the cortex (AUCs = 0.84–0.86) and spinal cord (AUCs = 0.76–0.86) holdout cohorts (**Fig. S17A, S17B**). However, improved predictive accuracy was seen when the seven-gene classifiers were applied to the HiSeq holdout cohort, with AUCs ranging from 0.86–0.91 (**Fig. S17C**), suggesting the seven-gene classifier may outperform the three-gene as the ‘strength’ of batch effects and confounding covariates increases. Collectively, our classification results demonstrate that the set of ALS-Ox marker genes established in this work can achieve appreciable stratification accuracy when predicting patient phenotype between regions of the central nervous system – with different cell type composition – or when using different instrumentation for quantification of gene expression.

## Discussion

Variable onset and progression of ALS has limited clinical trial success and slowed the development of effective therapeutics. This study builds on our previous study^5^ to show this large cohort of ALS patients^4^ can be stratified into three distinct molecular subtypes in both the postmortem cortex and spinal cord, with subtype-specific enrichment pathologies remaining mostly consistent. Our findings lend additional support to the biological role of these subtypes in disease progression and offers a promising foundation for development of more effective and personalized therapeutics. Conversely, our Cox survival analysis shows additional factors, other than molecular subtype, play a significant role in the variability in patient survival after adjustment for repeated patient measures and relevant covariates, but continue to support the relevance of these subtypes in the context of clinical heterogeneity. Drawing on other works, faster ALS progression has been broadly linked to neuroinflammation^23–26^, findings which are mostly consistent with the observed differences in subtype hazard and survival by the majority agreement method. However, additional work is needed to better understand how these phenotypes progress and interact with other factors like sex and disease comorbidity to drive variability in patient clinical parameters.

Comparing phenotypes assigned in the postmortem cortex and spinal cord, the lumbar region of the spinal cord was found to be most concordant with the cortical phenotype and most closely reflects the proportion of patients allocated to each subtype in the cortex^5^. To understand why, we consider work from Humphrey *et al.*^6^, which demonstrates cell type composition in the spinal cord of the same cohort acts as a major driver in observed gene expression differences. Keeping the limitations of bulk tissue RNA-seq in mind, the lumbar region of the spinal cord is reported to have the highest percentage of neuronal cells relative to glia^27^, allowing for a more detectable neuronal expression signature in the bulk tissue profile. Many of the altered pathways in the ALS-Ox subtype implicate neurons, leading to the conclusion that the higher percentage of these cell types in the lumbar region allows for improved stratification of the ALS cohort. As ALS patient stratification matures and clinical translation begins to take shape, the authors recommend biomarker sampling take place in the lumbar region of the spinal cord to reduce cell type composition influences. Generally, concordance between the frontal and medial motor cortex and the cervical and lumbar regions of the spinal cord are statistically significant, increasing the likelihood that patient subtypes persist throughout the central nervous system and influence the disease course.

In the ALS-Glia and ALS-TD spinal cord, many neuroinflammatory genes from the cortex^5^ were similarly elevated, likely reflecting cell type composition and stringent filtering of covariate-dependent gene expression. Enrichment analysis further supports shared phenotype themes between the two subtypes, suggesting the spinal cord may be less suited for stratification of ALS-Glia and ALS-TD patients, when using bulk tissue expression. Conversely, some of the most differentially expressed transcripts in each subtype broadly captured pathological themes observed in the postmortem cortex^5^. ALS-Glia samples maintain the most elevated expression of inflammatory genes, while ALS-TD samples primarily show the highest expression of non-coding transcripts, including pseudogenes, transcription factors, and long intergenic non-coding RNA. Further, expression of transcripts *MYL9*, *ST6GALNAC2*, and *TAGLN* were elevated in both the postmortem cortex and spinal cord of ALS-Glia patients, relative to the other subtypes and controls, providing a foundation for complete stratification using spinal cord expression. Interestingly, these marker genes were not directly involved in neuroinflammation and instead related to muscle contraction and protein glycosylation – indicating neuroinflammation is not a unique facet of the ALS-Glia spinal cord. We observe a further seven transcripts (‘ALS-Ox marker genes’) uniquely elevated in the postmortem cortex and spinal cord of ALS-Ox patients, supporting an ALS-Ox -specific pathology involving dysregulated and altered synaptic signaling. Identification of shared marker genes between the cortex and spinal cord lends strength to a generalized and coherent subtype presented at the patient level, although our unsupervised clustering and concordance analyses reveal the challenges surrounding the assignment of a single subtype to a patient. Again, additional work is necessary to address this gap, potentially including examination of time-dependent phenotype progression and association of subtype with other relevant clinical measures like MUNE, MRI imaging, dendritic density, electrophysiological recordings, and the qualitative ALSFRS-R score.

Demonstrating the utility of the ALS-Ox marker genes, we construct a variety of different machine learning classifiers that achieve impressive stratification accuracy in three unique holdout cohorts. While a similar analytical interpretation is obtained from the cortex and spinal cord validation cohorts, we specifically construct both to illustrate the global nature of elevated transcript expression and emphasize the capacity to use either region to predict the other. From a patient perspective, there is likely more benefit in the ability to use the spinal cord to predict the phenotype in the cortex – although as research grows the authors anticipate both directions may be relevant. The sequencing platform (HiSeq) holdout may best estimate predictive performance of our classifiers when applied to new patient cohorts with inconsistent batch effects and confounding factors. Lending strength to the biological relevance of the ALS-Ox marker genes, our classifiers continue to demonstrate high predictive accuracy when trained on NovaSeq samples and tested on HiSeq, and greatly outperform the ∼300 gene classifiers we developed previously^5^. Finally, the demonstrated ability to refine the set of seven ALS-Ox marker genes to achieve mostly equivalent predictive performances reduces barriers to clinical implementation and diagnostic burden. Looking forward, additional patient cohorts are needed to validate the utility of the ALS-Ox marker genes, including a consideration of expression from living individuals. The newfound ability to predict, with reasonable accuracy, the cortical phenotype from spinal tissue expression reduces the invasiveness of stratification procedures and provides an important foundation to validate the relevance of ALS-Ox marker genes *in vivo*.

## Methods

### Study approval

The NYGC ALS Consortium samples presented in this work were acquired through various IRB protocols from member sites and the Target ALS postmortem tissue core and transferred to the NYGC in accordance with all applicable foreign, domestic, federal, state, and local laws and regulations for processing, sequencing, and analyses^4^.

Postmortem brain tissues from cognitively normal individuals were obtained from the Mayo Clinical Florida Brain Bank. Diagnosis was independently ascertained by trained neurologists and neuropathologists upon neurological and pathological examinations, respectively. Written informed consent was given by all participants or authorized family members, and all protocols were approved by the IRB and ethics committee of the Mayo Clinic^4^.

### Data sources

GSE153960 contains RNA-seq data from 1659 tissue samples, spanning 11 regions of the CNS, from 439 patients with ALS, frontotemporal lobar degeneration (FTLD), or comorbidities for ALS-Alzheimer’s (ALS/AD) or ALS-FTLD. Patients were filtered such that only the individuals belonging to the groups ALS-TDP, ALS/FTLD, ALS/AD, and ALS-SOD1 were included in the ALS disease cohort. Patient samples were further filtered to consider the postmortem spinal cord exclusively, yielding 428 unique tissue transcriptomes from the cervical, thoracic, and lumbar regions. Tissue-matched control samples were obtained from the same publicly available dataset, totaling 91 tissue transcriptomes from 56 non-neurological control patients. Cohort demographics for this analysis are included in **Table S1**. Four samples that passed our inclusion criteria were excluded from the analysis due to consistent file transfer issues (SRR12166443, SRR12166526, SRR12166549, SRR12166553). Patient and sample IDs are de-identified and not known outside the research groups.

### Quantification

Quantification of gene expression was performed using STAR alignment^28^ and RSEM^29^, as detailed by Prudencio *et al*.^4^, and the processed gene count matrix was accessed directly from the GEO Accession (GSE153960). For quantification of transposable elements using SQuIRE, raw paired-end FASTQ files for all ALS and non-neurological control patient samples were downloaded from the European Bioinformatics Institute data repository (NCBI mirror) using Globus software. SQuIRE’s Fetch, Clean, Map, and Count functions were utilized as indicated^7^ to quantify locus-specific transposable elements. The expectation maximized ‘tot_counts’ values were selected as the estimate for sequencing reads attributed to each transposable element with gene locus resolution. The hg38 build was used during mapping, with default trim and EM parameters, and a read length of 100 or 125 base pairs depending on the sequencing platform specified. A scoring threshold of ≥ 99 was used to restrict the number of false positive TEs with few uniquely mapping reads. Stringent filtering was then applied to ensure all TEs included in the downstream analysis had at least one count for all available ALS samples (*n* = 428), resulting in 475 unique TE features (**Supplementary Data 1**).

### Clustering

Estimation of factorization rank was determined using the NMF package^30^ in R (Version 4.0.3, The R Foundation for Statistical Computing, Vienna, Austria)^31^. Quality measures were determined for ranks spanning 2 to 6, using 50 iterations at each rank and the default seeding method. The nsNMF (non-smooth non-negative matrix factorization) method was used for all NMF clustering.

Given previous work^6^, it was understood that cell type composition strongly influences bulk tissue expression in the spinal cord and used marker genes defined by the same study to remove these tissue-dependent features. Glial marker genes were obtained from Table S3 in *ref. 6*. Using DESeq2^8^, we further found a cumulative of 22,563 genes in the NovaSeq cohort and 17,804 genes in the HiSeq that were differentially expressed due to (i) sex, (ii) site of collection (NYGC versus Target ALS), (iii) RIN, and (iv) tissue region with an FDR adjusted *p*-value less than 0.05. After identifying dependent gene expression, patient transcriptomes were first subject to a variance stabilizing transformation, covariate-dependent genes were then removed, and filtering was applied such that the top 5,000 most variable genes (by median absolute deviation) were selected for clustering. Non-negative matrix factorization and visualization was performed in SAKE^10^ (Version 0.4.0). No samples were removed during the quality control step and further data transformations were not necessary. To robustly assign ALS sample subtype, 11 rounds of NMF clustering were performed in SAKE for both sequencing platform cohorts. A rank of three was used for each independent round of clustering, with 100 iterations per round, and the nsNMF algorithm. All software package versions have been detailed previously^5^.

### Enrichment

After each replicate of NMF clustering, gene and TE feature scores were calculated for all 5,000 MAD transcripts^32^. Feature scores were averaged across nsNMF clustering replicates and reordered. All features from both sequencing platform cohorts were combined, and after the removal of duplicates, 8163 transcripts remained for enrichment, corresponding to 5438 gene symbols.

For GSEA^13^, transcript expression was normalized to the DESeq2 median-of-ratios scale. Default parameters were maintained, aside from lowering the minimum gene set size to 5 and maximum to 150. We leveraged canonical pathways contained in the Reactome database^12^ and present pathway-level normalized enrichment scores for each ALS subtype. Non-neurological controls were specified as the reference level for subtype enrichment. To further support subtype-specific pathway enrichment observed in GSEA, we performed hypergeometric enrichment analysis using Enrichr^11^, the Reactome 2022 database, and our feature assignment approach detailed previously^5^. Enrichment *p*-values are determined by Fisher’s exact test, and presented as –log_10_ transformed values after FDR adjustment. The *p*-value heatmap is color-coded to indicate upregulation or downregulation relative to the other subtypes, and blank cells indicate an FDR adjusted *p*-value > 0.05.

### Clinical Parameters

The majority of patients in this cohort have more than one observation from the postmortem spinal cord. As a consequence, patient clinical parameters are considered using the majority agreement approach detailed previously^1,5^. In brief, patients were assigned a label only if there was a majority consensus among their cervical, thoracic, and lumbar samples, or if there was a single sample characterized, and labeled ‘Discordant’ in all other cases. Using this approach, differences in ALS patient survival were assessed using the Kaplan-Meier analysis^14^ with application of the log-rank statistical test. Subtype-level differences in age at symptom onset and death were analyzed by ANOVA with post hoc students’ t-tests (two-sided, unequal variance) and FDR *p*-value adjustment. Chi-squared tests of independence were applied to assess subtype-specificity for FTLD and Alzheimer’s comorbidity.

### Cox Proportional Hazard Regression

To address sample dependence due to repeat patient measures, we constructed a Cox proportional hazard regression model^16,18–20^ in R to assess multivariate contribution to patient survival. Sex, subtype, age at symptom onset, and disease group covariates are included as fixed effects and obtain hazard ratios from the exponential of the beta coefficient. Regression diagnostics show the proportional hazard assumption is met for all models and terms (**Figure S6**). By running regression in each tissue region independently, we bypass the need to incorporate patient-specific random effects in our proportional hazard model. The R function call for the proportional hazard regression model is provided in the **Supporting Information Text** and requires the ‘survival’ R library^18^. Model term *p*-values were calculated from the coefficient z-scores, while testing of the proportional hazard assumption at the covariate level was performed using the score test, with global *p* < 0.05 indicating time-dependent hazard and assumption violation^18,19^. All covariates in all models are observed to meet the assumption of having proportional hazards over the survival duration, excluding the disease group covariate in the lateral motor cortex model (*p* = 0.04).

### Concordance Analysis

Postmortem cortex subtype labels were previously determined^5^ and utilized in the present study to assess concordance with the molecular phenotype presented in the spinal cord of the same patients. Agreement is considered at the tissue-level rather than CNS level (i.e. cortex, spinal cord), to avoid sample dependence concerns with the majority agreement approach. Subtype discordance between the cortex and spinal is color-coded using a previous scheme from the hybrid subtyping analysis^5^, to inform which subtype label was more common, given the postmortem cortex observation. The *p*-values were estimated using bootstrapping, where distributions for the expected number of concordant patient samples were generated from 10,000 iterations, after adjusting sampling probabilities to reflect true observations from the cortex and spinal cord. Subtype re-sampling probabilities were adjusted separately for the postmortem cortex and spinal cord cohorts, with probabilities in the cortex set equal to 239/451, 84/451, and 128/451 for Ox, Glia, and TD subtypes respectively. The spinal cord probabilities were 139/428, 106/428, and 183/428, for Ox, Glia, and TD respectively. The same approach was used when considering concordance in the NovaSeq and HiSeq platforms independently, with all values presented in **Figure S3**. True concordant values were compared against the derived distributions for estimation of *p*-values assuming a one-tailed binomial distribution.

### Differential Expression

Differential transcript expression between ALS subtypes was considered using DESeq2^8^ with counts presented on the log_2_ scale, following size factor normalization (median-of-ratios). All patient samples were used to estimate size factors for normalization (n = 519 samples). A multifactor design equation was implemented, which included platform, site, RIN, tissue, and subtype covariates. Pairwise comparisons were performed using the contrast() argument, and FDR-adjusted *p*-values < 0.05 were considered to be significant. For presentation as a heatmap, transcript expression was z-score normalized, and observations that fell outside four standard deviations were adjusted to ±4 for plotting purposes only. FDR-adjusted *p*-values were –log_10_ transformed prior to plotting. Differential expression analysis from the cortex was determined previously and reused in the presentation of subtype marker genes^5^.

Given relevant work from Tam *et al.*^1^, Prudencio *et al*.^4^ and Humphrey *et al*.^6^ – which consider *TARDBP*, truncated stathmin-2, and cell type composition from the same cohort – we reexamine these features in the context of the stratified ALS cohort. We utilized DESeq2 to assess whether *TARDBP* expression was specific to a single subtype, and include all available observations from the postmortem cortex of the same patients for reference^5^. The normal length and truncated form of *STMN2* were determined previously by Prudencio *et al*. and provided by the NYGC ALS Consortium. Similarly, cell type composition – estimated from cell deconvolution – were previously determined by Humphrey *et al.*^6^ and are publicly available in the supplementary information.

### Classification

Classifiers to stratify ALS-Ox from all other patients (‘NotOx’) – including FTLD (*n*=42 cortex samples^5^) and non-neurological controls (*n*=93 cortex samples^5^ and *n*=91 spinal samples) – were developed using an 80/20 train/test split and three unique holdout cohorts comprised of (i) all cortical transcriptomes, (ii) all spinal cord transcriptomes, and (iii) all samples analyzed by HiSeq. 100-fold cross-validation was used to estimate F1 scores, with predictions made using the max distance metric and the first component. PLS-DA was performed using the ‘Mixomics’ library in R^21^.

Using the same train/test split, cross-validation, and holdout cohorts, additional classifiers were developed in Python (Version 3.9.10, Python Software Foundation, Wilmington, DE)^33^ using the scikit-learn framework^22^ (Version 1.3.0). Five different models were considered, which included k-nearest neighbors (KNN), linear discriminant analysis (LDA), multilayer perceptron (MLP), random forest (RF), and support vector machine classification (SVM). Default parameters were maintained unless otherwise noted. For the k-nearest neighbor classifier the number of neighbors was set to 8. For the SVM, a linear kernel was used with the regularization parameter, ‘C’ set to 0.025. Finally, the multilayer perceptron classifier was built using three hidden layers, with 100 ‘neurons’ comprising each hidden layer. The learning rate was set to 0.0001, while alpha was set equal to 1E-5.

## Data Availability

The raw RNA-seq data files used in this study are available in the NCBI Run Selector database under accession code PRJNA644618. The RSEM processed gene count matrix utilized in this study are available in the Gene Expression Omnibus database under accession code GSE153960. Transposable elements and processed RNA-seq count files are available as supplemental datasets or made publicly available at: https://figshare.com/authors/Jarrett_Eshima/13813720.

## Code Availability

All code developed and utilized in this analysis is available in the Barbara Smith Lab Github repository (https://github.com/BSmithLab/SpinalCordStratification).

## Acknowledgements

The authors would like to acknowledge The Target ALS Human Postmortem Tissue Core, New York Genome Center for Genomics of Neurogenerative Disease, Amyotrophic Lateral Sclerosis Association, Tow Foundation, and the patients and family members for supporting this analysis. The authors would like to thank Charles Pennington for his technical assistance and insight surrounding supercomputing architecture, SSH keys, and GitHub. J.E. is supported by the National Science Foundation, Graduate Research Fellowship (026257-001). John Fricks received financial support from the French government in the framework of the France 2030 program IdEx Université de Bordeaux.

## Notes

### Competing Interest Statement

The authors have declared no competing interest.

### Funding Statement

This study was funded by the Flinn Foundation (22-06449). J.E. is supported by the National Science Foundation, Graduate Research Fellowship (026257-001). J.F. received financial support from the French government in the framework of the France 2030 program IdEx Universite de Bordeaux

### Author Declarations

Original Study Approval: The NYGC ALS Consortium samples presented in this work were acquired through various IRB protocols from member sites and the Target ALS postmortem tissue core and transferred to the NYGC in accordance with all applicable foreign, domestic, federal, state, and local laws and regulations for processing, sequencing, and analyses. Postmortem brain tissues from cognitively normal individuals were obtained from the Mayo Clinical Florida Brain Bank. Diagnosis was independently ascertained by trained neurologists and neuropathologists upon neurological and pathological examinations, respectively. Written informed consent was given by all participants or authorized family members, and all protocols were approved by the IRB and ethics committee of the Mayo Clinic.

## References

[1] O.H. Tam, N.V. Rozhkov, R. Shaw, D. Kim, I. Hubbard, S. Fennessey, N. Propp, The NYGC ALS Consortium, D. Fagegaltier, B.T. Harris, L.W. Ostrow, H. Phatnani, J. Ravits, J. Dubnau, M.G. Hammell, Postmortem cortex samples identify distinct molecular subtypes of ALS: retrotransposon activation, oxidative stress, and activated glia. Cell Rep. 29, 1164–1177 (2019).

[2] E. Aronica, F. Baas, A. Iyer, A.L.M.A. ten Asbroek, G. Morello, S. Cavallaro, Molecular classification of amyotrophic lateral sclerosis by unsupervised clustering of gene expression in motor cortex. Neurobiol. Dis. 74, 359–376 (2015).

[3] G. Morello, M. Guarnaccia, A.G. Spampinato, S. Salomone, V. D’Agata, F.L. Conforti, E. Aronica, S. Cavallaro, Integrative multi-omic analysis identifies new drivers and pathways in molecularly distinct subtypes of ALS. Sci. Rep. 9, 1–8 (2019).

[4] M. Prudencio, J. Humphrey, S. Pickles, A. Brown, S.E. Hill, J.M. Kachergus, J. Shi, M.G. Heckman, M.R. Spiegel, C. Cook, Y. Song, M. Yue, L.M. Daughrity, Y. Carlomagno, K. Jansen-West, C. F. de Castro, M. DeTure, S. Koga, Y. Wang, P. Sivakumar, C. Bodo, A. Candalija, K. Talbot, B.T. Selvaraj, K. Burr, S. Chandran, J. Newcombe, T. Lashley, I. Hubbard, D. Catalano, D. Kim, N. Propp, S. Fennessey, NYGC ALS Consortium, D. Fagegaltier, H. Phatnani, M. Secrier, E.M.C. Fisher, B. Oskarsson, M. van Blitterswijk, R. Rademakers, N.R. Graff-Radford, B.F. Boeve, D.S. Knopman, R.C. Petersen, K.A. Josephs, E.A. Thompson, T. Raj, M. Ward, D.W. Dickson, T.F. Gendron, P. Fratta, L. Petrucelli, Truncated stathmin-2 is a marker of TDP-43 pathology in frontotemporal dementia. J. Clin. Investig. 130, e139741 (2020).

[5] J. Eshima, S.A. O’Connor, E. Marschall, NYGC ALS Consortium, R. Bowser, C.L. Plaisier, B.S. Smith, Molecular subtypes of ALS are associated with differences in patient prognosis. Nat. Commun., 14, 95 (2023).

[6] J. Humphrey, S. Venkatesh, R. Hasan, J.T. Herb, K. de Paiva Lopes, F. Küçükali, M. Byrska-Bishop, U.S. Evani, G. Narzisi, D. Fagegaltier, NYGC ALS Consortium, K. Sleegers, H. Phatnani, D.A. Knowles, P. Fratta, T. Raj, Integrative transcriptomic analysis of the amyotrophic lateral sclerosis spinal cord implicates glial activation and suggests new risk genes. Nat. Neurosci. 26, 150–162 (2023).

[7] W.R. Yang, D. Ardeljan, C.N. Pacyna, L.M. Payer, K.H. Burns, SQuIRE reveals locus-specific regulation of interspersed repeat expression. Nucleic acids Res. 47, e27–e27 (2019).

[8] M.I. Love, W. Huber, S. Anders, Moderated estimation of fold change and dispersion for RNA-seq data with DESeq2. Genome Biol. 15, 1–21 (2014).

[9] A. Pascual-Montano, J.M. Carazo, K. Kochi, D. Lehmann, R.D. Pascual-Marqui, Nonsmooth nonnegative matrix factorization (nsNMF). IEEE Trans. Pattern Anal. Mach. Intell. 28, 403–415 (2006).

[10] Y.J. Ho, N. Anaparthy, D. Molik, G. Mathew, T. Aicher, A. Patel, J. Hicks, M.G. Hammell, Single-cell RNA-seq analysis identifies markers of resistance to targeted BRAF inhibitors in melanoma cell populations. Genome Res. 28, 1353–1363 (2018).

[11] M.V. Kuleshov, M.R. Jones, A.D. Rouillard, N.F. Fernandez, Q. Duan, Z. Wang, S. Koplev, S.L. Jenkins, K.M. Jagodnik, A. Lachmann, M.G. McDermott, C.D. Monteiro, G.W. Gundersen, A. Ma’ayan, Enrichr: a comprehensive gene set enrichment analysis web server 2016 update. Nucleic acids Res. 44, W90–W97 (2016).

[12] B. Jassal, L. Matthews, G. Viteri, C. Gong, P. Lorente, A. Fabregat, K. Sidiropoulos, J. Cook, M. Gillespie, R. Haw, F. Loney, B. May, M. Milacic, K. Rothfels, C. Sevilla, V. Shamovsky, S. Shorser, T. Varusai, J. Weiser, G. Wu, L. Stein, H. Hermjakob, P. D’Eustachio, The reactome pathway knowledgebase. Nucleic acids Res. 48, D498–D503 (2020).

[13] A. Subramanian, P. Tamayo, V.K. Mootha, S. Mukherjee, B.L. Ebert, M.A. Gillette, A. Paulovich, S.L. Pomeroy, T.R. Golub, E.S. Lander, J.P. Mesirov, Gene set enrichment analysis: a knowledge-based approach for interpreting genome-wide expression profiles. Proc. Natl. Acad. Sci. U.S.A 102, 15545– 15550 (2005).

[14] E.L. Kaplan, P. Meier, Nonparametric estimation from incomplete observations. J. Am. Stat. Assoc. 53, 457–481 (1958).

[15] Y. Benjamini, Y. Hochberg, Controlling the false discovery rate: a practical and powerful approach to multiple testing. *J. R. Stat. Soc.*, B 57, 289–300 (1995).

[16] D.R. Cox, Regression models and life-tables. J. R. Stat. Soc., B 34, 187–202 (1972).

[17] P.C. Austin, A tutorial on multilevel survival analysis: methods, models and applications. International Statistical Review 85, 185–203 (2017).

[18] T.M. Therneau, T. Lumley, Package ‘survival’. R. Top. Doc. 128, 28–33 (2015).

[19] T.M. Therneau, P.M. Grambsch, *Modeling Survival Data: Extending the Cox Model* (Springer, New York, 2000).

[20] A. Kassambara, M. Kosinski, P. Biecek, S. Fabian, survminer: Drawing Survival Curves using ‘ggplot2’. (2017). Available from: https://CRAN.R-project.org/package=survminer

[21] F. Rohart, B. Gautier, A. Singh, K.A. Lê Cao, mixOmics: An R package for ‘omics feature selection and multiple data integration. PLoS Comput. Biol. 13, e1005752 (2017).

[22] F. Pedregosa, G. Varoquaux, A. Gramfort, V. Michel, B. Thirion, O. Grisel, M. Blondel, P. Prettenhofer, R. Weiss, V. Dubourg, J. Vanderplas, A. Passos, D. Cournapeau, M. Brucher, M. Perrot, E. Duchesnay, Scikit-learn: machine learning in Python. J. Mach. Learn. Res. 12, 2825–2830 (2011).

[23] D.R. Beers, J.S. Henkel, Q. Xiao, W. Zhao, J. Wang, A.A. Yen, L. Siklos, S.R. McKercher, S.H. Appel, Wild-type microglia extend survival in PU. 1 knockout mice with familial amyotrophic lateral sclerosis. Proc. Natl. Acad. Sci. U.S.A 103, 16021–16026 (2006).

[24] S. Boillée, K. Yamanaka, C.S. Lobsiger, N.G. Copeland, N.A. Jenkins, G. Kassiotis, G. Kollias, D.W. Cleveland, Onset and progression in inherited ALS determined by motor neurons and microglia. Science 312, 1389–1392 (2006).

[25] K. Yamanaka, S.J. Chun, S. Boillée, N. Fujimori-Tonou, H. Yamashita, D.H. Gutmann, R. Takahashi, H. Misawa, D.W. Cleveland, Astrocytes as determinants of disease progression in inherited amyotrophic lateral sclerosis. Nat. Neurosci. 11, 251–253 (2008).

[26] L. Vu, K. Garcia-Mansfield, A. Pompeiano, J. An, V. David-Dirgo, R. Sharma, V. Venugopal, H. Halait, G. Marcucci, Y.H. Kuo, L. Uechi, R.C. Rockne, P. Pirrotte, R. Bowser, Proteomics and mathematical modeling of longitudinal CSF differentiates fast versus slow ALS progression. Annals of clinical and translational neurology 10, 2025–2042 (2023).

[27] J. Bahney, C.S. von Bartheld, The cellular composition and glia–neuron ratio in the spinal cord of a human and a nonhuman primate: comparison with other species and brain regions. The Anatomical Record 301, 697–710 (2018).

[28] A. Dobin, C.A. Davis, F. Schlesinger, J. Drenkow, C. Zaleski, S. Jha, P. Batut, M. Chaisson, T.R. Gingeras, STAR: ultrafast universal RNA-seq aligner. Bioinformatics 29, 15–21 (2013).

[29] B. Li, C.N. Dewey, RSEM: accurate transcript quantification from RNA-Seq data with or without a reference genome. BMC Bioinforma. 12, 1–6 (2011).

[30] R. Gaujoux, C. Seoighe, A flexible R package for nonnegative matrix factorization. BMC bioinformatics 11, 1–9 (2010).

[31] R Core Team (2020). R: A language and environment for statistical computing. R Foundation for Statistical Computing, Vienna, Austria. URL https://www.R-project.org/.

[32] H. Kim, H. Park, Sparse non-negative matrix factorizations via alternating non-negativity-constrained least squares for microarray data analysis. Bioinformatics 23, 1495–1502 (2007).

[33] G. Van Rossum, F.L. Drake, Python 3 Reference Manual. (CreateSpace Independent Publishing Platform, Scotts Valley, CA, 2009).

## SI References

1. M. Prudencio, J. Humphrey, S. Pickles, A. Brown, S.E. Hill, J.M. Kachergus, J. Shi, M.G. Heckman, M.R. Spiegel, C. Cook, Y. Song, M. Yue, L.M. Daughrity, Y. Carlomagno, K. Jansen-West, C. F. de Castro, M. DeTure, S. Koga, Y. Wang, P. Sivakumar, C. Bodo, A. Candalija, K. Talbot, B.T. Selvaraj, K. Burr, S. Chandran, J. Newcombe, T. Lashley, I. Hubbard, D. Catalano, D. Kim, N. Propp, S. Fennessey, NYGC ALS Consortium, D. Fagegaltier, H. Phatnani, M. Secrier, E.M.C. Fisher, B. Oskarsson, M. van Blitterswijk, R. Rademakers, N.R. Graff-Radford, B.F. Boeve, D.S. Knopman, R.C. Petersen, K.A. Josephs, E.A. Thompson, T. Raj, M. Ward, D.W. Dickson, T.F. Gendron, P. Fratta, L. Petrucelli, Truncated stathmin-2 is a marker of TDP-43 pathology in frontotemporal dementia. J. Clin. Investig. 130, e139741 (2020).

2. J. Eshima, S.A. O’Connor, E. Marschall, NYGC ALS Consortium, R. Bowser, C.L. Plaisier, B.S. Smith, Molecular subtypes of ALS are associated with differences in patient prognosis. Nat. Commun., 14, 95 (2023).

3. O.H. Tam, N.V. Rozhkov, R. Shaw, D. Kim, I. Hubbard, S. Fennessey, N. Propp, The NYGC ALS Consortium, D. Fagegaltier, B.T. Harris, L.W. Ostrow, H. Phatnani, J. Ravits, J. Dubnau, M.G. Hammell, Postmortem cortex samples identify distinct molecular subtypes of ALS: retrotransposon activation, oxidative stress, and activated glia. Cell Rep. 29, 1164–1177 (2019).

4. J. Humphrey, S. Venkatesh, R. Hasan, J.T. Herb, K. de Paiva Lopes, F. Küçükali, M. Byrska-Bishop, U.S. Evani, G. Narzisi, D. Fagegaltier, NYGC ALS Consortium, K. Sleegers, H. Phatnani, D.A. Knowles, P. Fratta, T. Raj, Integrative transcriptomic analysis of the amyotrophic lateral sclerosis spinal cord implicates glial activation and suggests new risk genes. Nat. Neurosci. 26, 150–162 (2023).

5. X. Wang, J. Park, K. Susztak, N.R. Zhang, M. Li, Bulk tissue cell type deconvolution with multi-subject single-cell expression reference. Nat. Commun. 10, 380 (2019).

6. H. Mathys, J. Davila-Velderrain, Z. Peng, F. Gao, S. Mohammadi, J.Z. Young, M. Menon, L. He, F. Abdurrob, X. Jiang, A.J. Martorell, R.M. Ransohoff, B.P. Hafler, D.A. Bennett, M. Kellis, L. Tsai, Single-cell transcriptomic analysis of Alzheimer’s disease. Nature 570, 332–337 (2019).

7. E.L. Kaplan, P. Meier, Nonparametric estimation from incomplete observations. J. Am. Stat. Assoc. 53, 457–481 (1958).

8. Y. Benjamini, Y. Hochberg, Controlling the false discovery rate: a practical and powerful approach to multiple testing. *J. R. Stat. Soc.*, B 57, 289–300 (1995).

9. T.M. Therneau, T. Lumley, Package ‘survival’. R. Top. Doc. 128, 28–33 (2015).

10. S. Park, D.J. Hendry, Reassessing Schoenfeld residual tests of proportional hazards in political science event history analyses. American Journal of Political Science 59, 1072–1087 (2015).

11. M.I. Love, W. Huber, S. Anders, Moderated estimation of fold change and dispersion for RNA-seq data with DESeq2. Genome Biol. 15, 1–21 (2014).

12. W.R. Yang, D. Ardeljan, C.N. Pacyna, L.M. Payer, K.H. Burns, SQuIRE reveals locus-specific regulation of interspersed repeat expression. Nucleic acids Res. 47, e27–e27 (2019).

