## Supplementary Information for "Elevated expression of *B4GALT6*, *GABRA1*, *GAD2*, *GLRA3*, *HTR2A*, *PCSK1*, and *SLC17A6* are postmortem markers for the ALS-Ox subtype"

Science Advances

AAAS

Jarrett Eshima *et al.*

**This PDF file includes:**

Supplementary Text
Figs. S1 to S17
Tables S1
References (1 to 12)

**Other Supplementary Materials for this manuscript include the following:**

Data S1 to S6

**Supporting Information Text**

**R Call for Cox Proportional Hazards Regression Model.**

```
> coxph(Surv(time) ~ SubjectSex + AgeOnset + disease_group + Subtype, data=CNS_MEM, model = T)
```

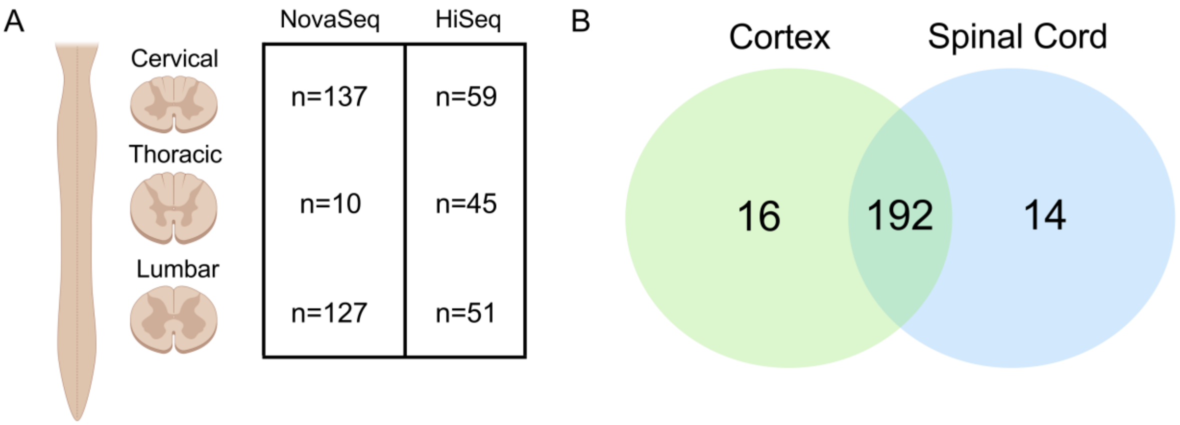


**Fig. S1. Overview of GSE153960 spinal cord transcriptomes considered in this study.** (**A**) Sample number corresponding to each region of the spinal cord analyzed by Prudencio et al.[1] on the NovaSeq and HiSeq sequencing platforms. (**B**) Patient level analysis showing a high degree of overlap between the spinal cord ALS cohort and cortex cohort analyzed previously[2].

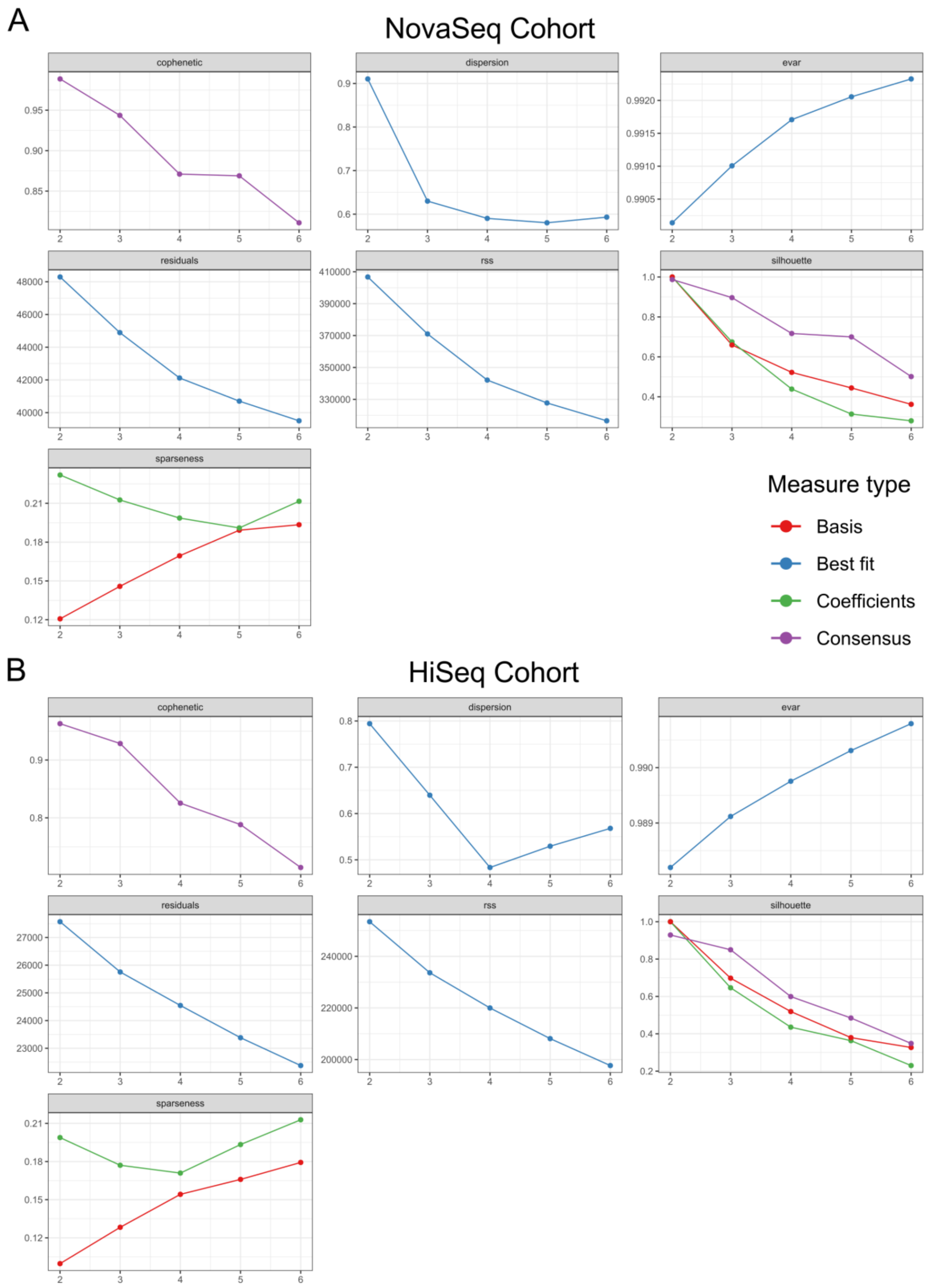


**Fig. S2. Factorization rank for nsNMF clustering.** Factorization rank was determined by estimating clustering metrics from 50 iterations in (**A**) the NovaSeq cohort and (**B**) the HiSeq cohort for ranks spanning 2 to 6 (*x-axis*). In both cohorts, the non-smooth non-negative matrix factorization (nsNMF) method was used.

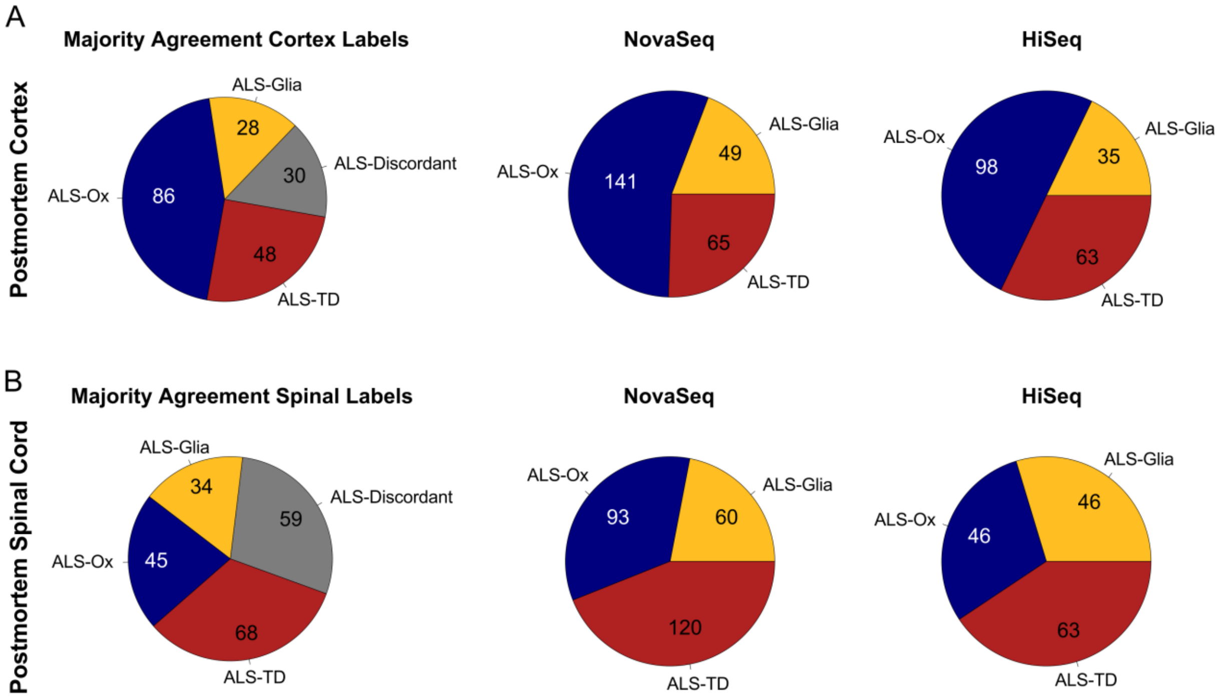


**Fig. S3. Generalized comparison of subtypes assigned to the ALS postmortem cortex and spinal cord.** Pie charts showing patient level subtypes using the majority agreement approach described previously[2,3] in the (**A**) postmortem cortex and (**B**) the postmortem spinal cord. A larger fraction of patients was found to be "Discordant" or "ALS-TD" in the spinal cord as compared to the cortex, potentially reflecting cell type composition differences in each tissue region. When accounting for sequencing platform in the assignment of patient subtype at the individual sample level, the NovaSeq cohort more closely mirrors the subtype proportion in the postmortem cortex. Yet, in both cases, the ALS-Ox subtype was not the most common, again pointing to cell composition effects given work from Humphrey's *et al*.[4]

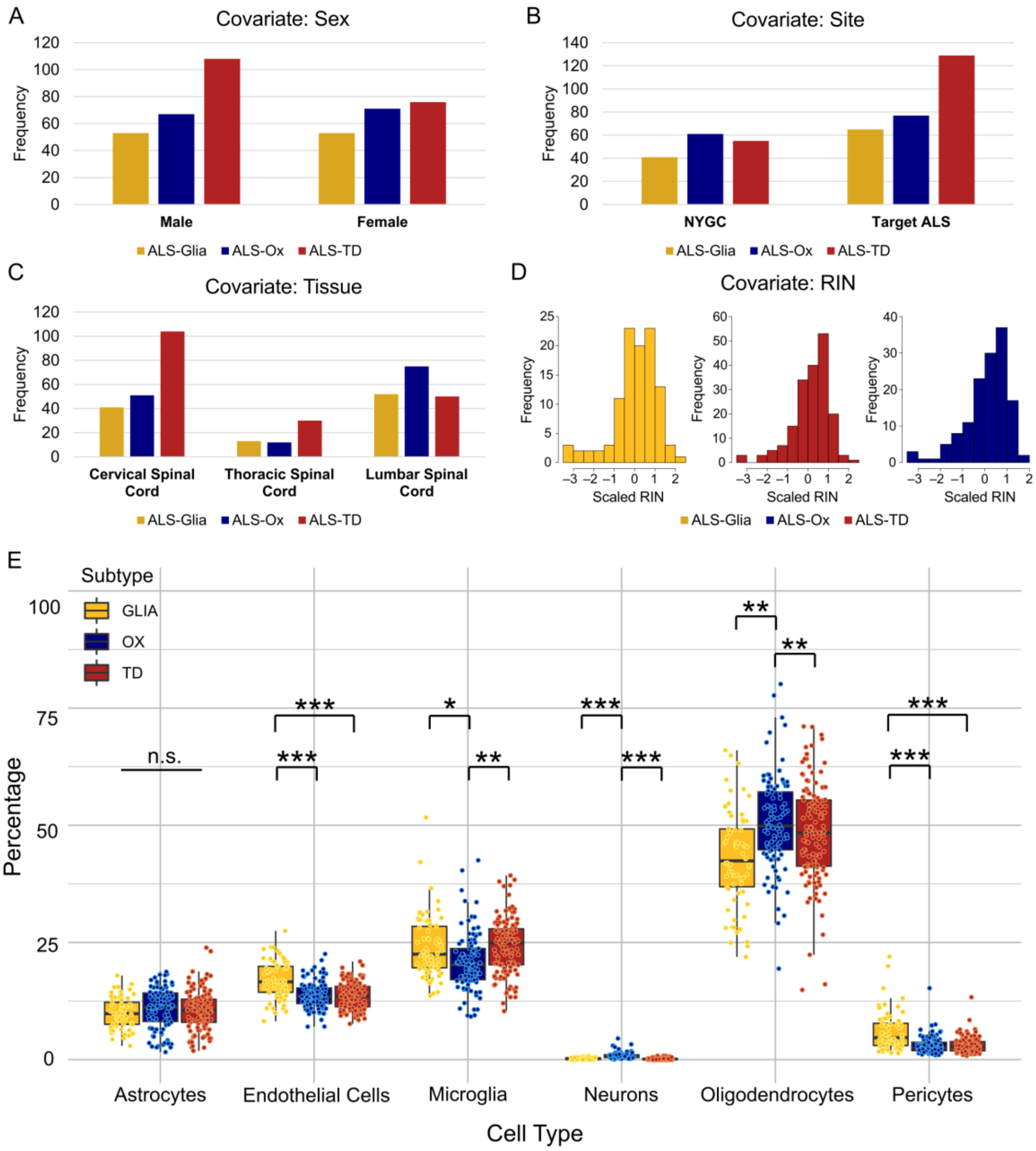


**Fig. S4. Diagnostic plots show spinal cord subtype is partially influenced by multiple covariates after dependent gene removal.** Covariate diagnostic plots following assignment of sample subtype using non-smooth non-negative matrix factorization showing (**A**) sex, (**B**) site of collection, (**C**) tissue region, and (**D**) RIN. While scaled RIN does not appear to have a strong effect on the assigned subtype, the remaining covariates are seen to influence subtype despite removal of covariate-dependent genes using differential expression. Strictly considering this spinal cord cohort, the NYGC site of collection/processing and the lumbar tissue region appear more robust to covariate-dependent gene expression. (**E**) Estimated cell type fractions in the postmortem spinal cord (n = 293 unique transcriptomes, 137 cervical, 36 thoracic, 120 lumbar), considered in the context of the ALS subtypes. Estimates were previously calculated by Humphrey *et al.*[4] using the MuSiC algorithm[5] with reference single-nucleus RNA-seq data from Mathys *et al.*[6] Significant differences in cell type percentages were assessed using a two-sided Wilcoxon rank sum test with Bonferroni $p$-value adjustment. Adjusted $p$-values are denoted using the following scheme: *** $p < 1E\text{-}5$; ** $p < 0.001$; * $p < 0.05$. Pairwise comparisons not depicted have an adjusted $p$-value > 0.05.

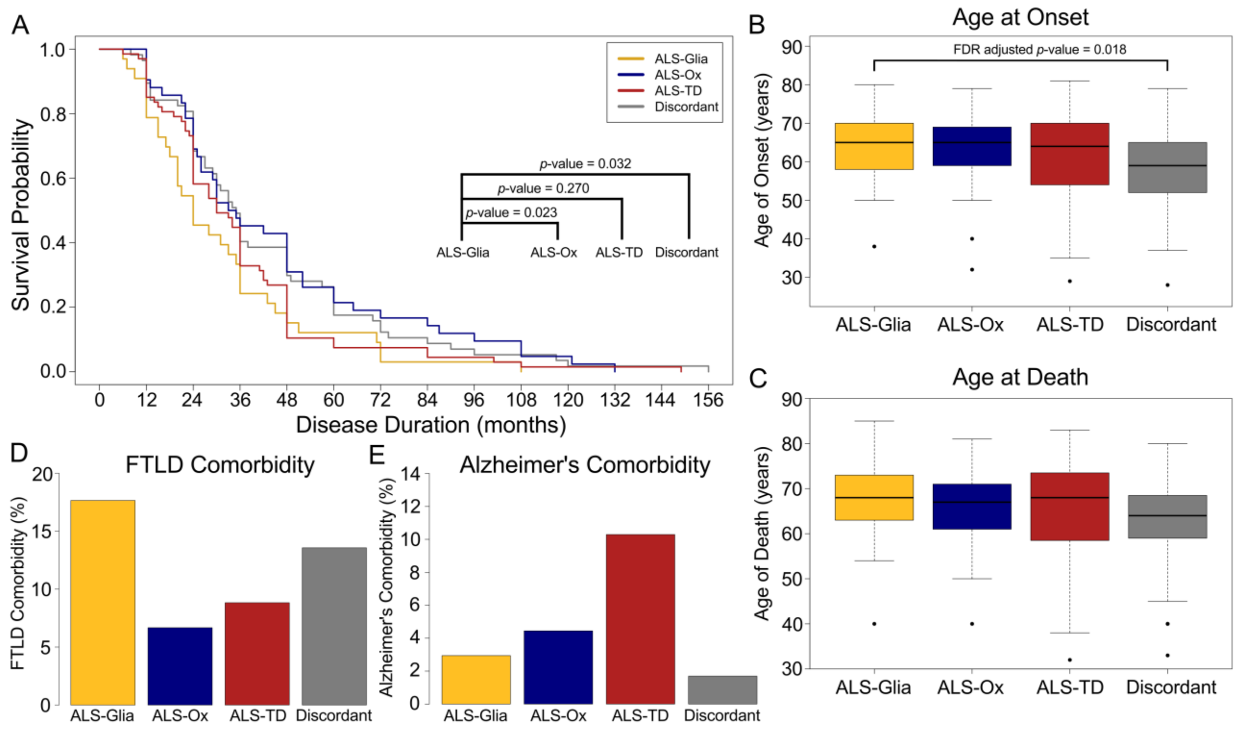


**Fig. S5. Survival and clinical parameters analysis.** (**A**) Kaplan-Meier survival analysis[7] using patient subtypes (*n*=206) defined by spinal cord transcriptomes. Subtypes were assigned if the majority of available tissue regions were concordant, otherwise the patients were assigned to the 'Discordant' group. The ALS-Glia subtype is observed to have a significantly shorter survival duration when compared to the ALS-Ox and Discordant groups. (**B**) Age of onset (*n*=206) and (**C**) age at death (*n*=206) are presented as boxplots for each subtype. T-tests with a false discovery rate[8] correction were applied, and the Glia subtype was seen to have a significantly later age of onset as compared to the Discordant group. Comorbidity for (**D**) FTLD and (**E**) Alzheimer's disease are presented as bar plots. Chi-squared tests of independence were performed and neither FTLD ($p$ = 0.38) nor Alzheimer's ($p$ = 0.15) comorbidity was seen to be associated with ALS subtype.

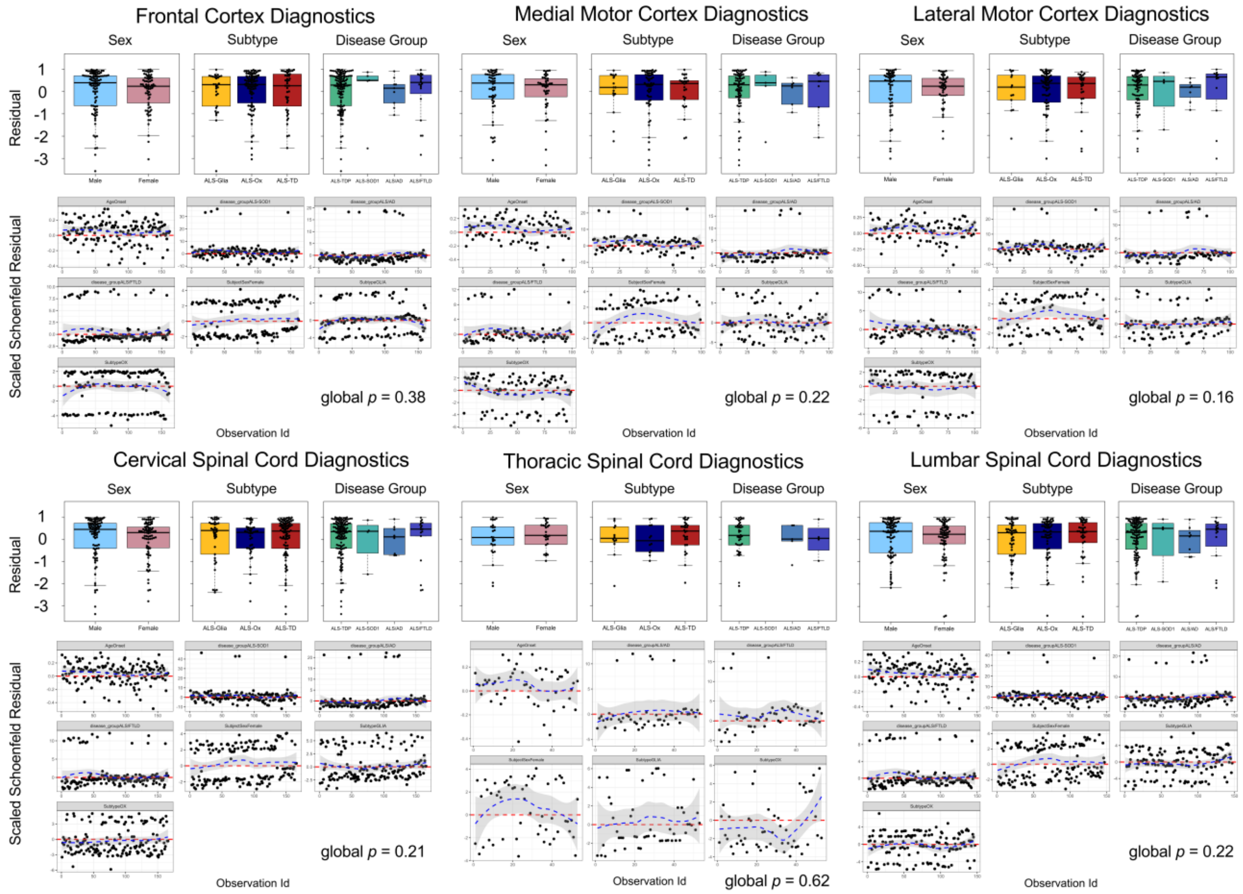


**Fig. S6. Cox proportional hazard regression diagnostics.** Sample level observations from the postmortem cortex and spinal cord were separated by tissue region and utilized to construct Cox proportional hazard regression models. Sex, disease group, age of onset, and subtype were included as model covariates, yielding a total of 728 observations without missing data in the six Cox models shown. For each model constructed, residuals are plotted by covariate, and generally show weak or null dependency on the variable level. To assess adherence to the proportional hazard model assumption, scaled Schoenfeld residual plots are shown for each covariate level, with score test *p*-values[9] determined using the ‘km’ time transformation[10]. All covariates are seen to meet the assumption of having proportional hazards over the survival duration, excluding the disease group covariate in the lateral motor cortex model ($p = 0.04$).

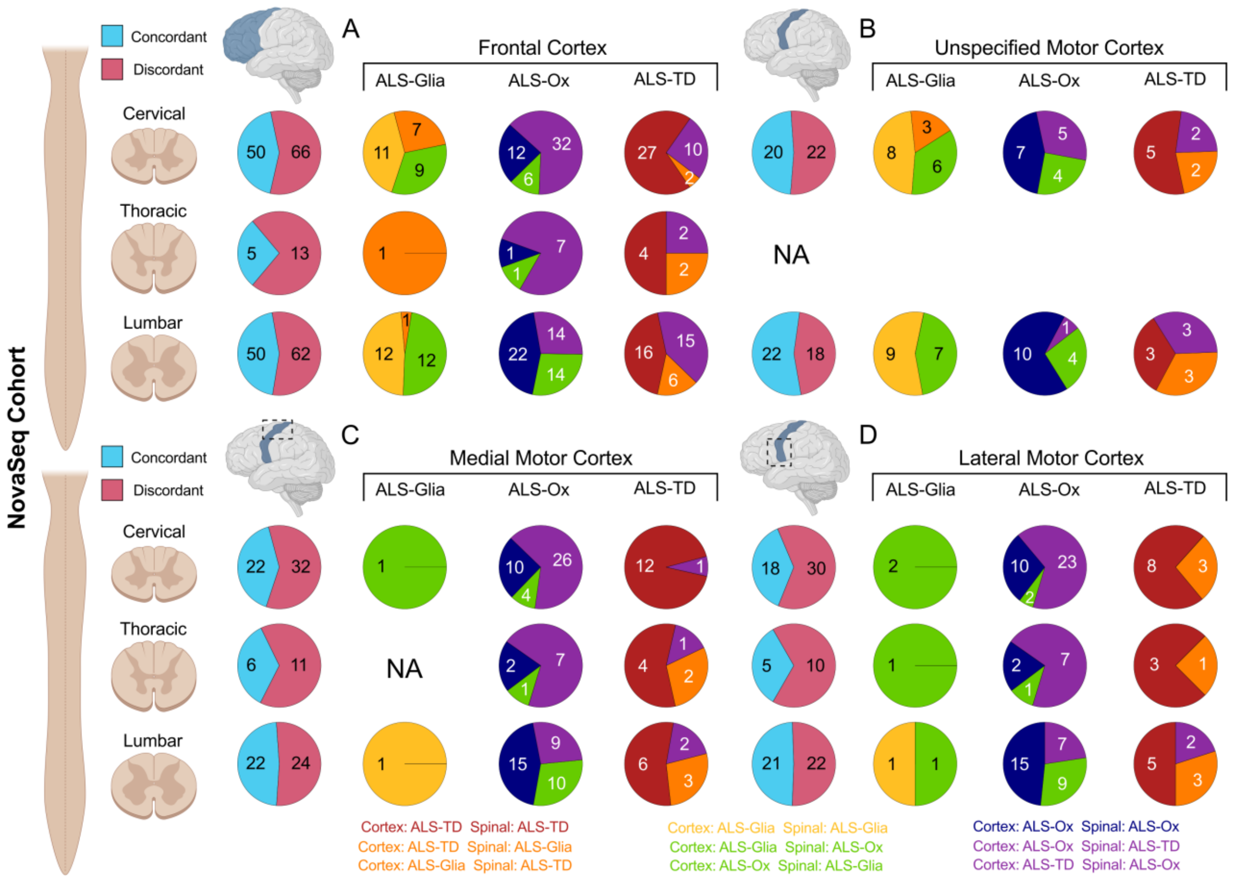


**Fig. S7. Tissue specific concordance between the postmortem cortex and spinal cord in the NovaSeq cohort.** Agreement between the subtype assigned to the cervical, thoracic, and lumbar regions (*rows*) and (**A**) the frontal cortex, (**B**) the unspecified motor cortex, (**C**) the medial motor cortex, and (**D**) the lateral motor cortex – for all available NovaSeq spinal cord samples. Pie charts are first presented as an aggregate of all paired tissue samples (light blue and pink) and in a subtype-specific manner. Concordance at the subtype level has been color coded to indicate agreement (gold, navy, and maroon) or disagreement (orange, green, and purple) between the two tissue regions compared. No patients assigned ALS-TD in the medial motor cortex had a corresponding thoracic spinal cord sample in this cohort. There were no intra-patient tissue pairings between the thoracic spinal cord and unspecified motor cortex in this cohort.

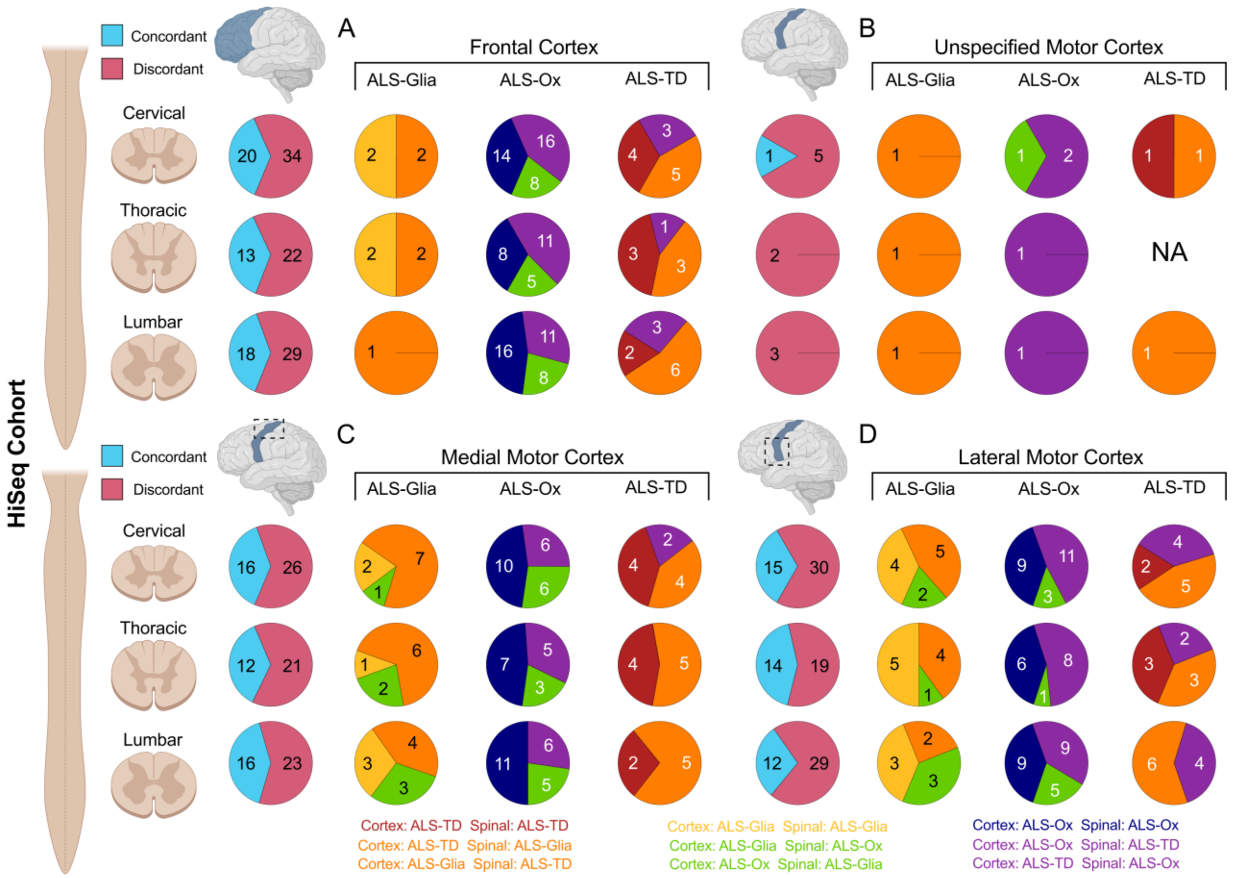


**Fig. S8. Tissue specific concordance between the postmortem cortex and spinal cord in the HiSeq cohort.** Agreement between the subtype assigned to the cervical, thoracic, and lumbar regions (*rows*) and (**A**) the frontal cortex, (**B**) the unspecified motor cortex, (**C**) the medial motor cortex, and (**D**) the lateral motor cortex – for all available HiSeq spinal cord samples. Pie charts are first presented as an aggregate of all paired tissue samples (light blue and pink) and in a subtype-specific manner. Concordance at the subtype level has been color coded to indicate agreement (gold, navy, and maroon) or disagreement (orange, green, and purple) between the two tissue regions compared. No patients assigned ALS-TD in the unspecified motor cortex had a corresponding thoracic spinal cord sample in this cohort.

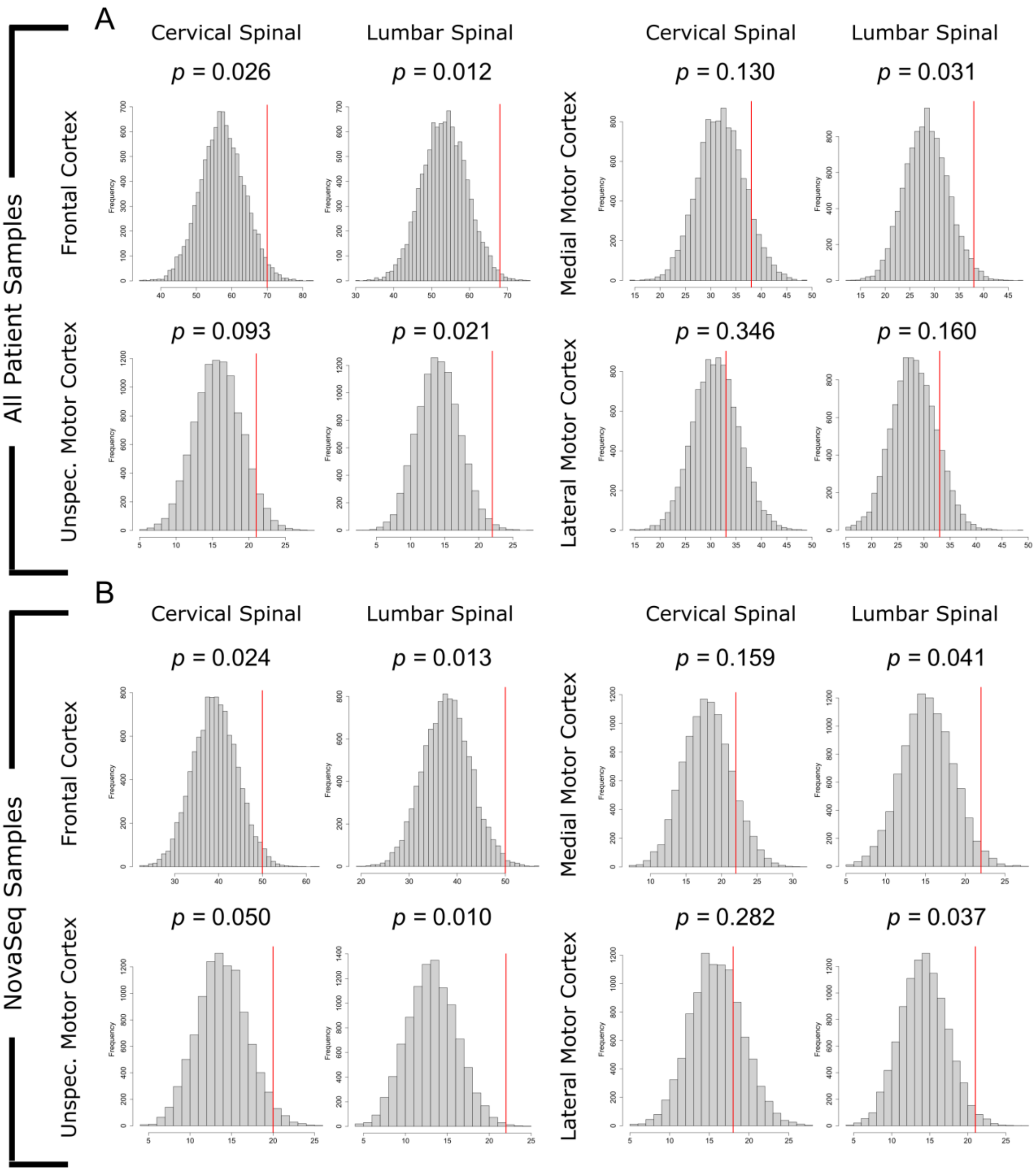


**Fig. S9. Histograms showing bootstrap-derived concordance *p*-values.** Concordance histograms are shown for each region of the postmortem cortex when compared to the cervical and lumbar regions of the spinal cord. The red line indicates the true number of concordant samples observed for a given cortex – spinal tissue pairing. *P*-value estimates from a one-tailed binomial distribution are provided for (A) all patient samples and (B) the NovaSeq subset exclusively. The NovaSeq platform generally outperforms the HiSeq, with no tissue pairings having statistical significance in the HiSeq subset. No significant agreement was observed in the thoracic spinal cord in any cohort.

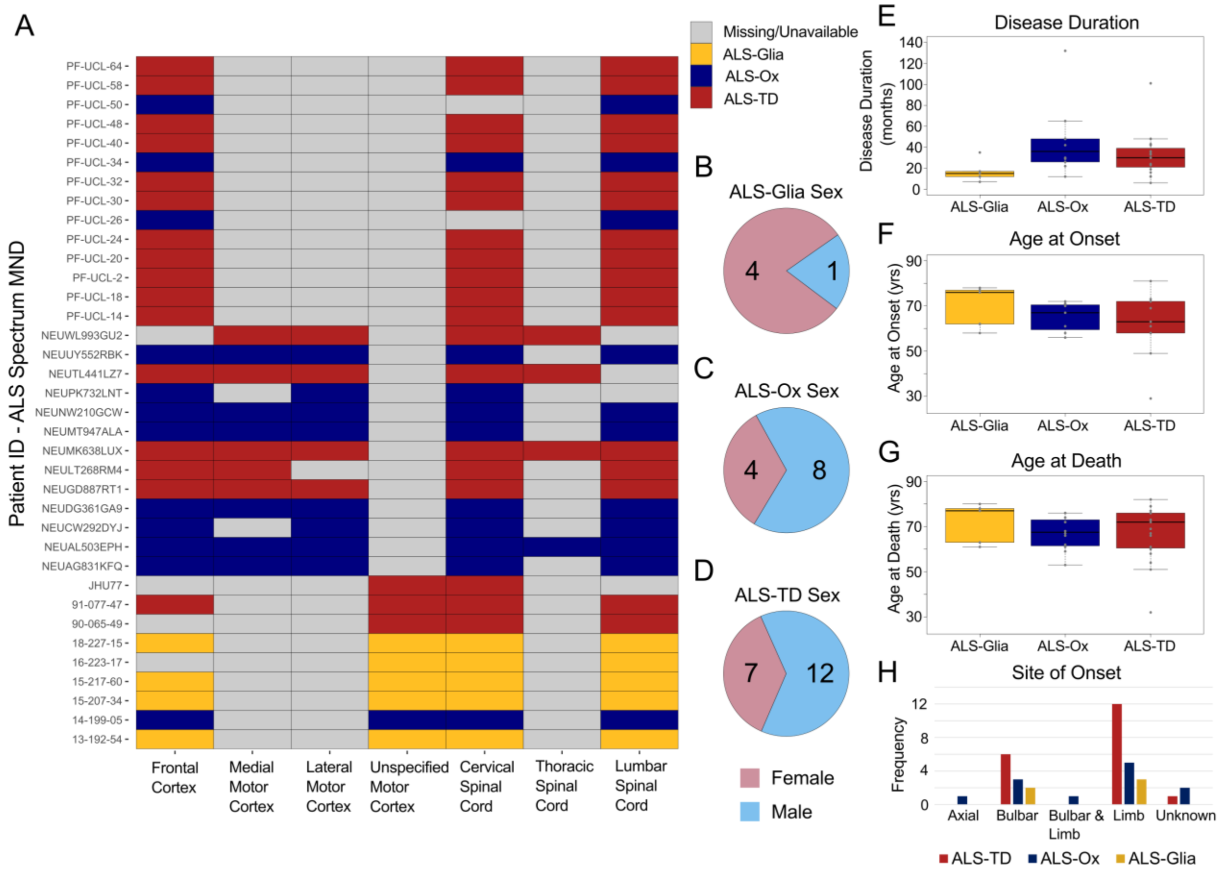


**Fig. S10. ALS patients with subtype concordance throughout the central nervous system.** Multiple patients demonstrated perfect concordance across all available tissue transcriptomes and are presented using publicly available de-identified IDs. (**A**) Heatmap showing subtype assignment to each region of the cortex and spinal cord considered in this study. Gray cells correspond to unavailable or not applicable tissue transcriptomes. Patients without observations from both the postmortem cortex and spinal cord are excluded from this plot, but are available in **Supplementary Dataset 4**. Patient sex is presented for (**B–D**) each ALS subtype. Interestingly, patients concordant for the ALS-Glia subtype are primarily female, potentially indicating sex-dependent differences in the presentation of disease phenotype. Clinical parameters for concordant patients are plotted as boxplots, and show (E) disease duration, (F) age at onset, (G) age at death, and (H) site of symptom onset. Statistical tests were not performed due to limited patient number.

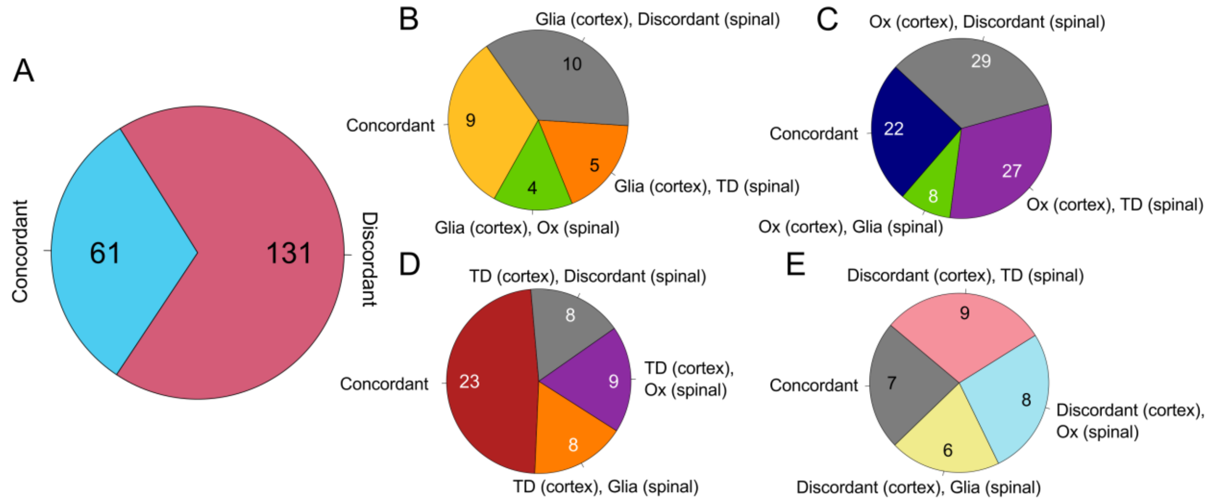


**Fig. S11. Concordance between the postmortem cortex and spinal cord using the majority agreement approach.** A meta level analysis comparing subtype assigned in the cortex with the spinal cord using the majority agreement approach – in which patients were assigned a subtype if a single sample was available or by majority if two or more samples were available. Patient subtype was assigned in the cortex and spinal cord independently. (**A**) The majority (68.2%) of patient samples did not show concordance between the postmortem cortex and spinal cord when using the majority agreement approach – indicating this method is not the optimal way to manage repeat patient sampling. (**B**) For patients that presented as ALS-Glia in the cortex, ~32% of individuals were assigned the same subtype in their spinal cord by majority agreement. Discordant was the most common subtype assigned, likely reflecting limitations due to cell type composition of the spinal cord. (**C**) Patients with the oxidative stress phenotype in the cortex demonstrated similar concordance, with ~25% of patients assigned the same subtype in their spinal cord. Encouragingly, very few patients classified as ALS-Ox in the cortex were assigned ALS-Glia in their spinal cord, suggesting cell type composition does not act as a confounding factor in the expression of ALS-Ox marker genes in the spinal cord but weakens the detectable signal. (**D**) Patients presenting as ALS-TD in their cortex showed the highest concordance with the spinal cord (~48%), but likely reflects bias towards this subtype in the spinal cord transcriptomes. Interestingly, this bias towards ALS-TD in the spinal cord does not appear to be due to RIN, given that the mean RIN was 6.14 for all cortex samples yet 6.50 for all samples from the spinal cord. (**E**) Patients initially showing discordance for their postmortem cortex subtype are reassigned to ALS subtypes with roughly the same frequency. The concordance label in this figure indicates both the postmortem cortex and spinal cord presented as “Discordant” by the majority agreement method.

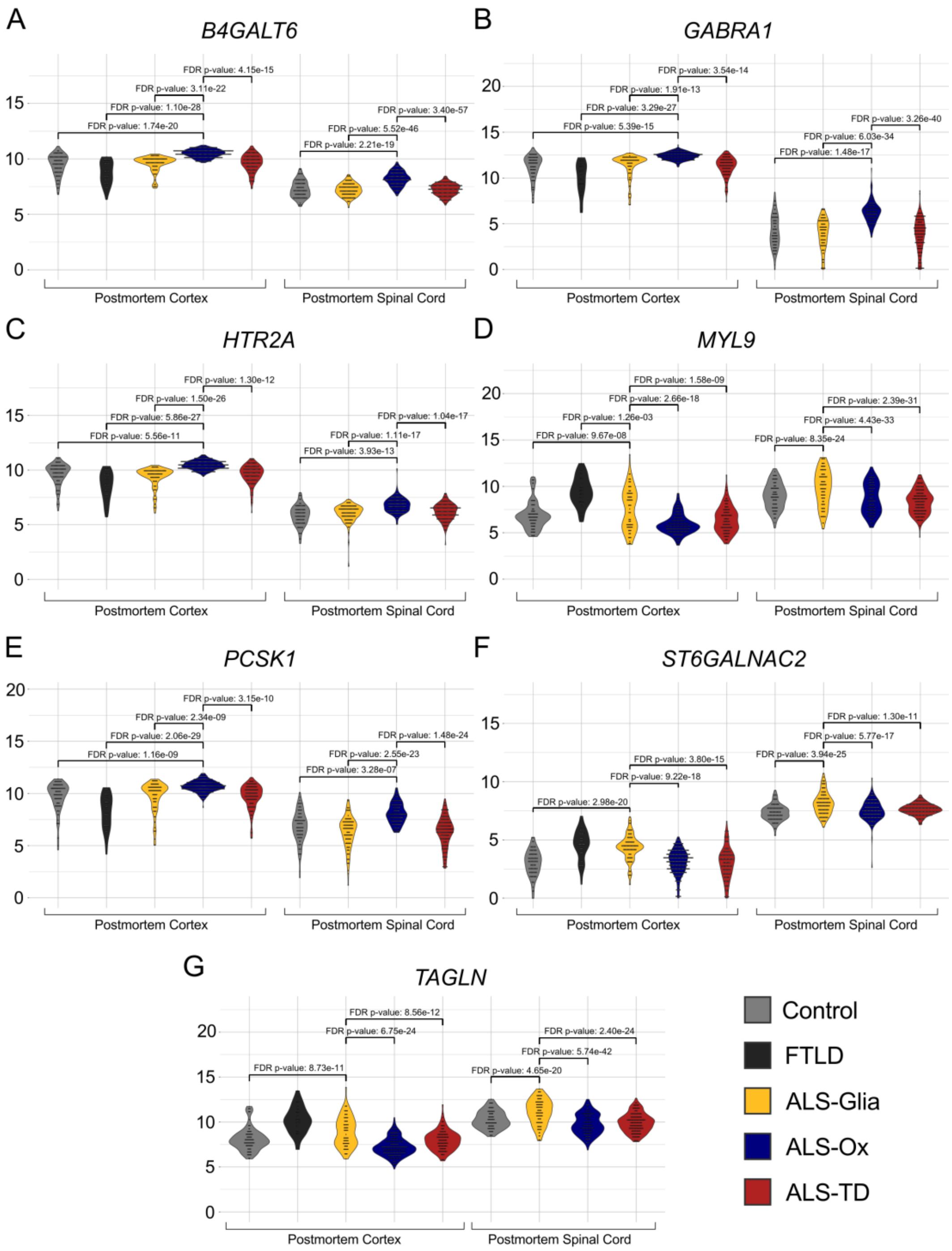


**Fig. S12. Subtype-specific marker genes in the postmortem cortex and spinal cord.** ALS-Ox and ALS-Glia marker genes show coherent expression throughout the central nervous system. Expression is separated both by subtype and CNS region[2] for (**A**) *B4GALT6*, (**B**) *GABRA1*, (**C**) *HTR2A*, (**D**) *MYL9*, (**E**) *PCSK1*, (**F**) *ST6GALNAC2*, and (**G**) *TAGLN*. All counts are presented on the $\log_2$ median-of-ratio scale[11]. All differential expression *p*-values are FDR adjusted. ALS-Ox marker genes include *B4GALT6, GABRA1, GAD2, GLRA3, HTR2A, PCSK1,* and *SLC17A6*. ALS-Glia marker genes include *MYL9*, *ST6GALNAC2*, and *TAGLN* although expression of these genes is less specific for ALS-Glia and are likely more susceptible to cell composition differences in the spinal cord.

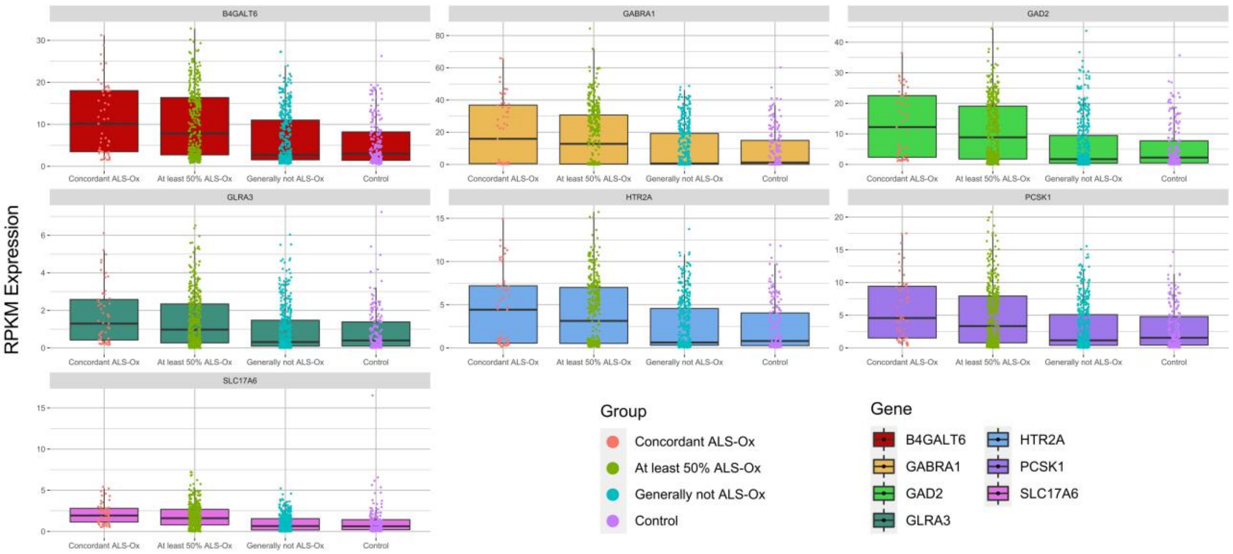


**Fig. S13. RPKM normalized ALS-Ox marker gene expression.** Sample-level expression of ALS-Ox marker genes after grouping by ALS-Ox percentage, calculated by taking the number of intra-patient samples defined as ALS-Ox divided by the total number of samples from the patient. Transcript expression is normalized by library size to the RPKM scale. Concordant ALS-Ox patients were defined as ALS-Ox in all available tissue samples, while the ‘generally not ALS-Ox’ category is defined as less than 50% of samples classified as ALS-Ox. A total of 53 unique samples were included in the ‘Concordant ALS-Ox’ category, 393 samples in the ‘at least 50% ALS-Ox’, 410 samples in ‘generally not ALS-Ox’, and 184 control samples – corresponding to 206 ALS patients and 88 non-neurological controls.

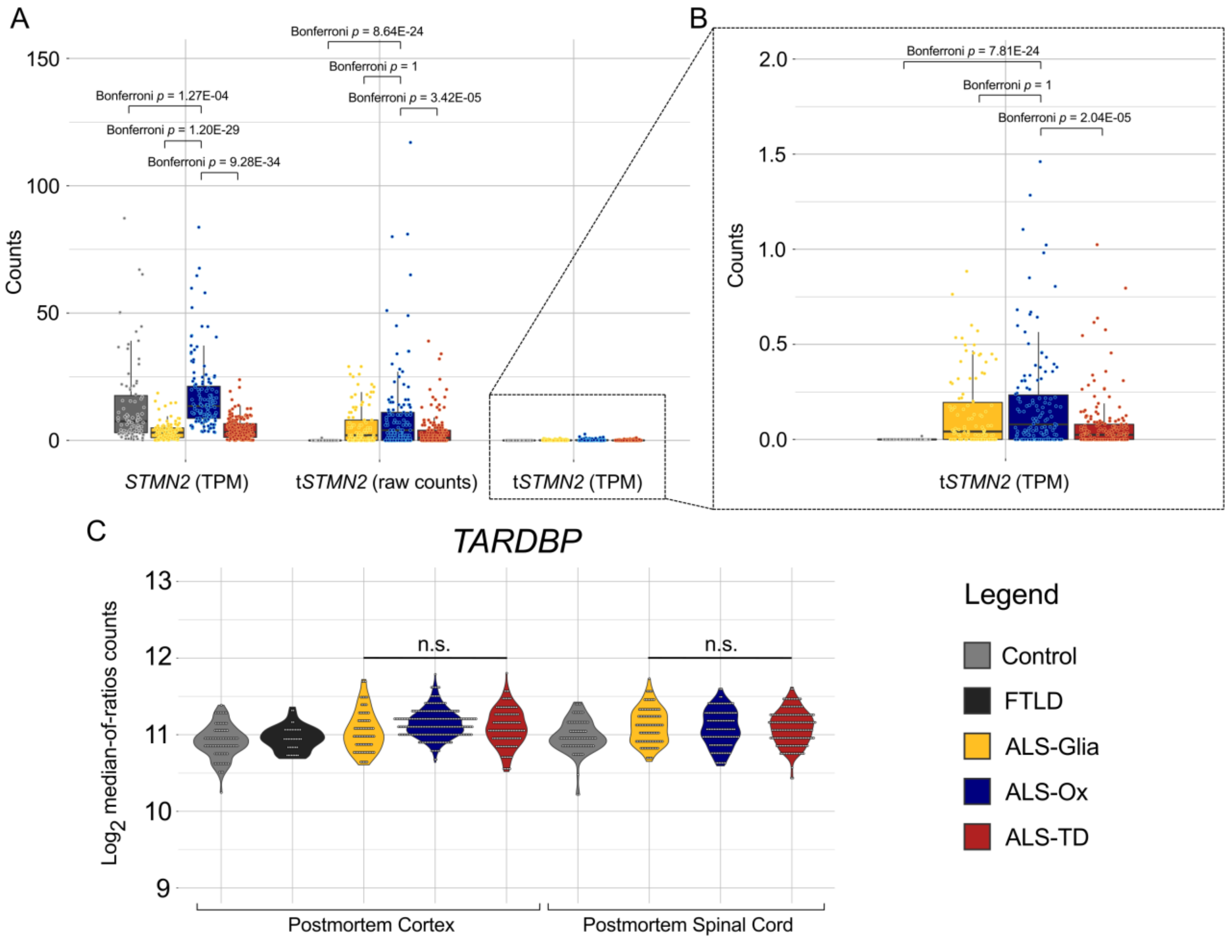


**Fig. S14. Expression of *STMN2*, truncated *STMN2*, and TARDBP.** (**A**) A two-sided Mann-Whitney U test was used to assess statistical significance in *STMN2* and truncated *STMN2* expression on both TPM scale and raw count scale. After adjusting p-values for multiple hypothesis testing using the Bonferroni method, truncated STMN2 expression was elevated in the postmortem spinal cord of ALS-Ox patients when compared to the ALS-TD subtype on both count scales, further supporting phenotypic differences between ALS-Ox and ALS-TD patients. These finds are somewhat surprising, given ALS-TD pathology appears more closely linked to transcription as compared to ALS-Ox. (**B**) Truncated STMN2 counts on the TPM scale are replotted for visual clarity. No statistically significant differences in the expression of STMN2 or truncated STMN2 are observed between ALS-Glia and ALS-TD subtypes. (**C**) Expression of transcript *TARDBP*, encoding ALS disease-associated protein TDP-43, is presented to show transcript level differences are not observed in the postmortem spinal cord, as well as the cortex[2]. These findings further support TDP-43 pathology occurring at the protein level rather than transcript level.

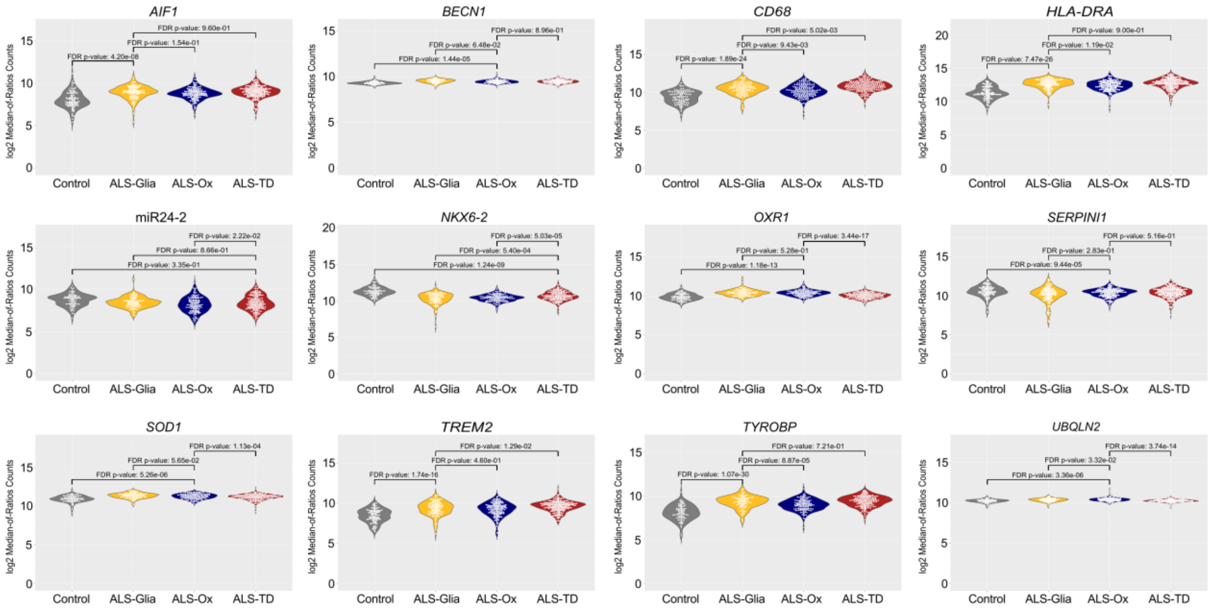


**Fig. S15. The neuroinflammatory subtype (ALS-Glia) is obscured in the ALS spinal cord.** Transcripts found to stratify this ALS cohort using the postmortem cortex[2] are reconsidered in the spinal cord. ALS-Glia cortex transcripts *AIF1*, *CD68*, *HLA-DRA*, *TREM2*, and *TYROBP* show weak or non-significant differences in expression compared to the other two subtypes in the spinal cord. Genes associated with proteotoxic and oxidative stress are elevated in the cortex of ALS-Ox patients but not in the spinal cord, seen in the expression of *BECN1*, *OXR1*, *SERPINI1*, *SOD1*, and *UBQLN2* yet tissue composition at the cellular level may partially explain these differences. *NKX6-2* but not miR24-2, both associated with the regulation of transcription, showed weak but consistent upregulation in the cortex and spinal cord of ALS-TD patients.

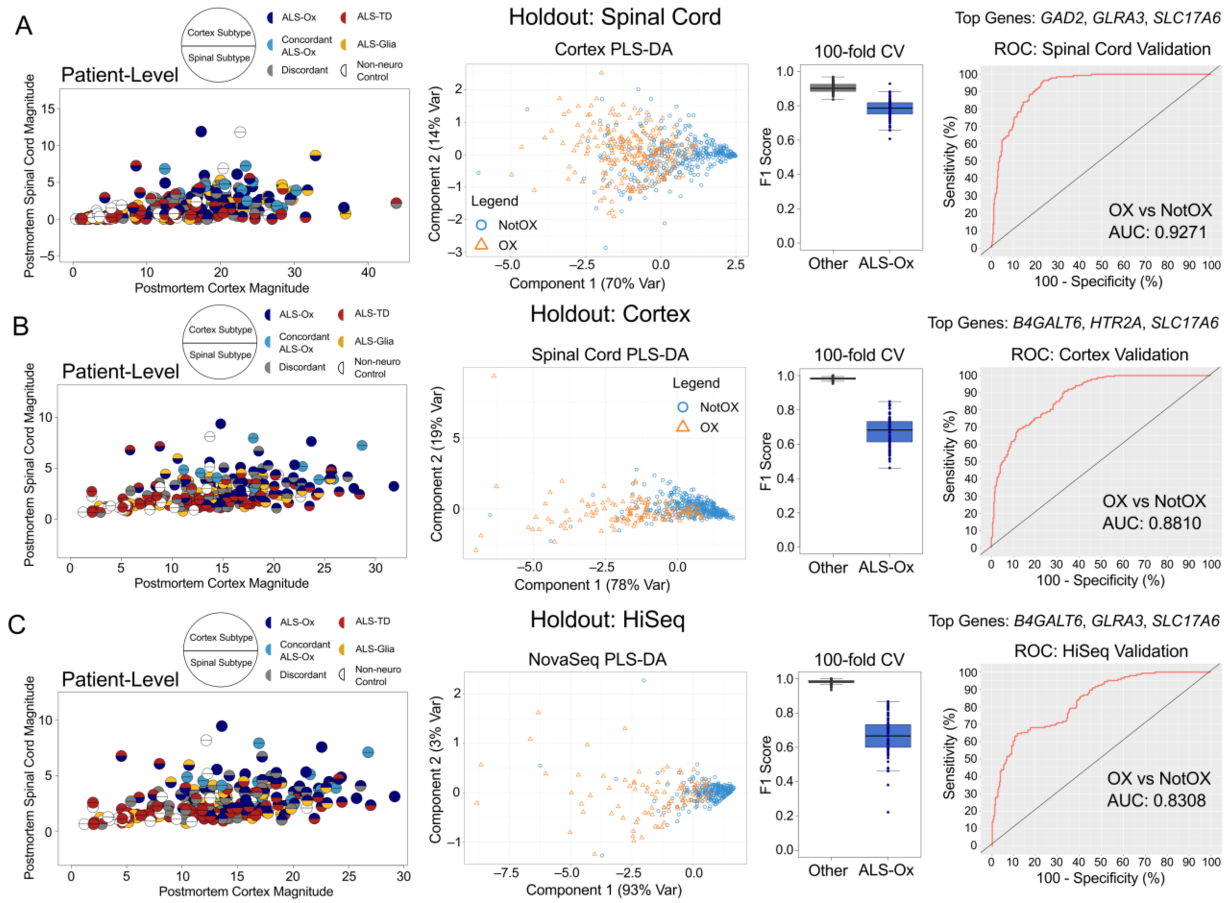


**Fig. S16. Three-gene PLS-DA classifiers for ALS-Ox patients.** Partial least squares discriminate analysis with expression of transcripts normalized to the RPKM scale using library size and transcript length estimates from GRCh38.p12. In each case, visualization of patients is first performed using ALS-Ox marker genes, taking the mean RPKM expression magnitude – for the three-gene combination – from each available tissue sample. Majority assigned subtype is color-coded with the postmortem cortex and spinal cord presented in the upper and lower half circles, respectively. The PLS-DA classifier was then trained and tested using an 80/20 split of (**A**) all postmortem cortex samples, and validated using all spinal cord samples, (**B**) all postmortem spinal cord samples, and validated on the cortex holdout, and (**C**) all NovaSeq samples, and validated on the HiSeq holdout. Following PLS-DA, the training cohort is plotted using the first two components. Test cohort F1 metrics are presented as boxplots for 100 rounds of cross validation predicting ALS-Ox against all other samples, including non-neurological controls (‘Other’). Lastly, ROC plots showing application of the PLS-DA classifier to each of the three holdout cohorts. The top gene combination is provided for each holdout cohort, and the *B4GALT6*, *GLRA3*, *SLC17A6* trio was found to have the highest average AUC across all three holdout datasets.

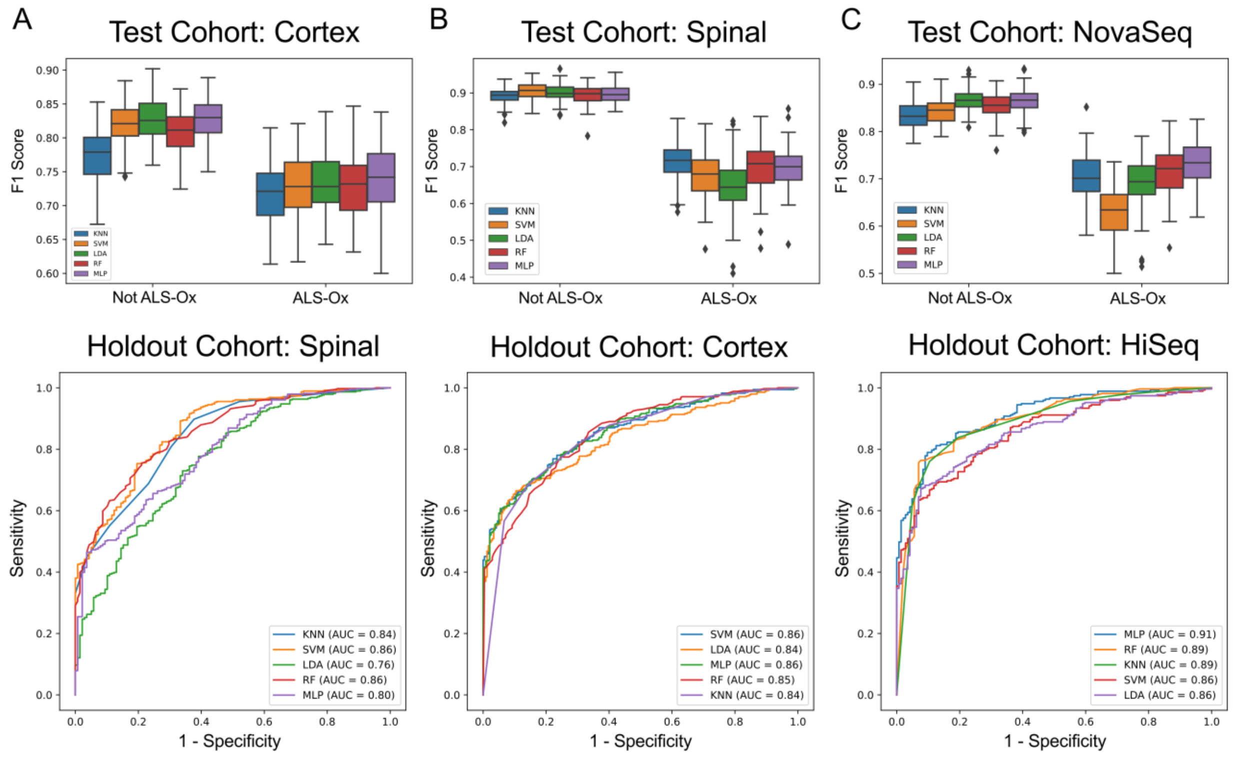


**Fig. S17. Supervised machine learning classifiers using all seven ALS-Ox marker genes.** Five different classifiers were constructed, using RPKM normalized expression of all seven ALS-Ox marker genes. F1 scores from 100 rounds of cross validation are presented as boxplots, for each classifier considered. F1 scores are separated by class level (ALS-Ox and 'Not ALS-Ox'). A combined total of 1,104 ALS and control samples are considered, with *n*=377 (~34%) assigned the ALS-Ox label. Classifiers were constructed and applied to three different holdout cohorts comprised of (**A**) all postmortem spinal cord samples (*n*=519), (**B**) all postmortem cortex samples (*n*=585), and (**C**) all samples analyzed by HiSeq (*n*=415). ROC plots are presented second, for each classifier, and show sensitivity vs 1-specificity metrics when applied to the specified holdout cohort.

| Cohort Demographics<br>A-L: Axial and Limb Onset<br>B-L: Bulbar and Limb Onset<br>A-B: Axial and Bulbar Onset | ALS Spectrum<br>(n = 206) | Healthy Control Donors<br>(n = 56) |
|---|---|---|
| Sex | | |
| Female | 97 (47.1%) | 31 (55.4%) |
| Male | 109 (52.9%) | 25 (44.6%) |
| Tissue Site | n = 428 | n = 91 |
| Cervical Spinal Cord | 195 (45.6%) | 40 (44.0%) |
| Thoracic Spinal Cord | 55 (12.9%) | 9 (9.9%) |
| Lumbar Spinal Cord | 178 (41.6%) | 42 (46.2%) |
| ALS Subtype | | NA |
| ALS-Glia | 34 (16.5%) | – |
| ALS-TD | 68 (33.0%) | – |
| ALS-Ox | 45 (21.8%) | – |
| ALS-Discordant | 59 (28.6%) | – |
| Disease Duration (months) | | NA |
| ALS-Glia | 31.3 ± 3.97 | – |
| ALS-TD | 36.0 ± 3.01 | – |
| ALS-Ox | 45.6 ± 4.87 | – |
| ALS-Discordant | 43.2 ± 3.98 | – |
| Age of Onset (years) | | NA |
| ALS-Glia | 64.7 ± 1.50 | – |
| ALS-TD | 61.6 ± 1.58 | – |
| ALS-Ox | 62.6 ± 1.63 | – |
| ALS-Discordant | 58.4 ± 1.41 | – |
| Site of Onset | | NA |
| ALS-Glia | A-B: 1; A-L: 1; Bulbar: 8; B-L: 1 Limb: 23 | – |
| ALS-TD | A-L: 1; Bulbar: 21; B-L: 1; Limb: 37; Unknown: 8 | – |
| ALS-Ox | Axial: 2; Bulbar: 12; B-L: 2; Limb: 26; Generalized: 1; Unknown: 2 | – |
| ALS-Discordant | Axial: 1; A-B: 1; Bulbar: 16; B-L: 1 Limb: 39; Unknown: 1 | – |
| Age of Death (years) | | 63.8 ± 2.16 * |
| ALS-Glia | 67.4 ± 1.48 | – |
| ALS-TD | 65.5 ± 1.36 | – |
| ALS-Ox | 66.1 ± 1.15 | – |
| ALS-Discordant | 62.7 ± 1.29 | – |
| FTLD Comorbidity | 23/206 (11.2%) | NA |
| ALS-Glia | 6/34 (17.6%) | – |
| ALS-TD | 6/68 (8.8%) | – |
| ALS-Ox | 3/45 (6.7%) | – |
| ALS-Discordant | 8/59 (13.6%) | – |

**Table S1. Cohort Demographics.** ALS patients and healthy controls considered in this analysis. Disease duration, age of onset, and age of death statistics are provided as mean ± standard error. *Two non-neurological control donors had an age of death listed as “90 or Older”. An estimate of 90 years was used for all samples listed as such.

**Dataset S1 (separate file).** Quantification of transposable elements in the spinal cord of all ALS and non-neurological control patient samples considered in this study using SQuIRE[12]. Filtering was applied to ensure at least one count was observed in all ALS patient samples, leaving a total of 475 unique transposable elements.

**Dataset S2 (separate file).** Patient and sample level metadata, including subtype assigned to all available tissue samples from the postmortem cortex and spinal cord.

**Dataset S3 (separate file).** Phenotype file used for the construction of the Cox proportional hazard regression models.

**Dataset S4 (separate file).** Metadata corresponding to the highly concordant patients presented in supplementary figure 10, and additional patients presenting a coherent subtype in all tissue samples from *either* the postmortem cortex or postmortem spinal cord. This information may be useful in defining reference expression for each phenotype and/or in follow-up studies comparing phenotypic differences between each subtype.

**Dataset S5 (separate file).** Differential gene expression results comparing subtypes in a pairwise manner.

**Dataset S6 (separate file).** RPKM normalized expression matrices and phenotype information for all patient samples (n = 1104) considered during classification analysis. The following splice variants from GRCh38.p12 were used as the estimate for the gene length: *B4GALT6* - ENST00000237019.11; *GABRA1* - ENST00000393943.10; *GAD2* - ENST00000259271.7; *GLRA3* - ENST00000274093.8; *HTR2A* - ENST00000378688.8; *MYL9* - ENST00000279022.7; *PCSK1* - ENST00000311106.8; *SLC17A6* - ENST00000263160.3; *ST6GALNAC2* - ENST00000225276.9; *TAGLN* - ENST00000278968.10.

### SI References


1. Prudencio, M. et al. Truncated stathmin-2 is a marker of TDP-43 pathology in frontotemporal dementia. *J. Clin. Investig.* **130**, e139741 (2020).
2. Eshima, J. et al. Molecular subtypes of ALS are associated with differences in patient prognosis. *Nature Communications*, **14**, 95 (2023).
3. Tam, O. H. et al. Postmortem cortex samples identify distinct molecular subtypes of ALS: retrotransposon activation, oxidative stress, and activated glia. *Cell Rep.* **29**, 1164–1177 (2019).
4. Humphrey, J. et al. Integrative transcriptomic analysis of the amyotrophic lateral sclerosis spinal cord implicates glial activation and suggests new risk genes. *Nature Neuroscience*, 1–13 (2023).
5. Wang X, Park J, Susztak K, Zhang NR, Li M. Bulk tissue cell type deconvolution with multi-subject single-cell expression reference. Nature communications. 2019 Jan 22;10(1):380.
6. Mathys H, Davila-Velderrain J, Peng Z, Gao F, Mohammadi S, Young JZ, Menon M, He L, Abdurrob F, Jiang X, Martorell AJ. Single-cell transcriptomic analysis of Alzheimer's disease. Nature. 2019 Jun 20;570(7761):332-7.
7. Kaplan, E. L. & Meier, P. Nonparametric estimation from incomplete observations. *J. Am. Stat. Assoc.* **53**, 457–481 (1958).
8. Benjamini, Y. & Hochberg Y. Controlling the false discovery rate: a practical and powerful approach to multiple testing. *Journal of the Royal statistical society: series B (Methodological)* **57**, 289–300 (1995).
9. Therneau, T. M. & Lumley, T. Package 'survival'. *R. Top. Doc.* **128**, 28–33 (2015).
10. Park S, Hendry DJ. Reassessing Schoenfeld residual tests of proportional hazards in political science event history analyses. American Journal of Political Science. 2015 Oct;59(4):1072-87.
11. Love, M. I., Huber, W. & Anders, S. Moderated estimation of fold change and dispersion for RNA-seq data with DESeq2. *Genome Biol.* **15**, 1–21 (2014).
12. Yang, W. R., Ardeljan, D., Pacyna, C. N., Payer, L. M. & Burns, K. H. SQuIRE reveals locus-specific regulation of interspersed repeat expression. *Nucleic acids Res.* **47**, e27–e27 (2019).
